# A Federated Learning-based Optic Disc and Cup Segmentation Model for Glaucoma Monitoring In Color Fundus Photographs

**DOI:** 10.64898/2025.12.02.25341485

**Authors:** Sudhanshu Shrivastava, Upasana Thakuria, Scott Kinder, Giacomo Nebbia, Nazlee Zebardast, Sally L. Baxter, Benjamin Xu, Saif Aldeen Aldeen Alryalat, Malik Kahook, Jayashree Kalpathy-Cramer, Praveer Singh

## Abstract

**Importance:** Glaucoma, a leading cause of blindness worldwide, depends on accurate optic nerve head assessment, particularly optic disc and cup segmentation, for diagnosis and monitoring. Deep learning (DL) models can automate these measurements, but models trained on smaller, site-specific datasets often fail to generalize. While larger, multi-site datasets help, data privacy concerns limit centralized training.

**Objective:** To evaluate a federated learning (FL) framework with site-specific fine-tuning for optic disc and cup segmentation, aiming to match central model performance while preserving privacy and improving generalizability.

**Design:** Comparative evaluation of three different approaches: (1) a central model trained on multi-site data, (2) site-specific local model training (3) standard FL models, against an FL with site-specific fine-tuning.

**Setting:** Multicenter study incorporating nine publicly available datasets, representing varied clinical environments, populations, and imaging protocols.

**Participants:** 5,550 color fundus photographs from at least 917 individuals across nine datasets includingboth routine care and research sources from 7 countries.

**Exposures:** Optic disc and cup segmentationin color fundus photographs using training with local model, central model, standard FL, and FL with site-specific fine-tuning..

**Main Outcomes and Measures:** Segmentation accuracy measured by Dice score. Comparisons were labeled as performance “wins” or “losses” based on statistically significant differences via Wilcoxon signed-rank test (P < 0.05).

**Results:** Site-specific fine-tuning of FL with site-specific fine tuning matched central model performance for cup segmentation across all sites (9/9) and for disc segmentation in most sites (7/9). Compared with site-specific local models, it preserved within-site performance (cup: 9/9; disc: 5/9) while substantially improving cross-site generalizability, achieving significant gains in 54.2% (39/72) of disc and 25.0% (18/72) of cup external-site evaluations, with no significant losses. Compared to standard FL pipelines, site-specific fine-tuning improved performance by 52% for disc and 26% for cup.

**Conclusions and Relevance:** Site-specific fine-tuning within an FL framework effectively personalizes generalized models to local data distributions, achieving central-level performance without data sharing and enhancing cross-site robustness. This approach enables privacy-preserving, scalable AI deployment across heterogeneous clinical settings for reproducible and generalizable glaucoma assessment

**KEY POINTS:** *Question:* How can we train an AI model to segment the optic cup and disc across multiple sites without sharing data, yet achieve performance comparable to a central model trained on pooled datasets?

*Findings:* In this federated learning (FL) study of 5,550 fundus photographs from nine sites, a site-specific fine-tuning FL strategy matched the central model’s performance and outperformed other standard FL techniques, with notable gains in cross-site generalizability.

*Meaning:* Site-specific fine-tuning effectively personalizes FL models to local data distributions, combining data privacy with robust, generalizable performance.

## INTRODUCTION

Glaucoma, a major global cause of irreversible blindness, affects 3.54% of the population aged 40-80 and is projected to impact 111.8 million people by 2040^1–3^. Early detection is crucial as the condition progresses silently, often resulting in significant vision loss if untreated^3–5^. A potential indicator of glaucoma severity is the vertical disc to cup ratio, measuring the ratio between the vertical diameter of the disc versus that of the cup, with ratios ≥0.6 suggestive of glaucoma^6–15^. Thus, optic disc and cup segmentation methods might be beneficial for detecting and monitoring glaucoma progression^1,16–18^. Given its clinical significance, various techniques have been developed to directly estimate the cup-to-disc ratio^19^ or segment both structures^20^.

Over the past decade, Deep Learning (DL) has significantly advanced automatic image analysis across various fields, such as radiology^21,22^, histopathology^23,24^, and ophthalmology^25–27^. Developing effective DL models requires large and diverse datasets^28–30^ for high accuracy and robustness, explaining the substantial size of datasets used in non-medical applications^30^. Such larger datasets, common in non-medical applications, are more challenging to compile in medical imaging due to privacy laws like the General Data Protection Regulation (GDPR) in Europe^31,32^ and the Health Insurance Portability and Accountability Act (HIPAA) in the USA^33^, and the difficulties in data aggregation across institutions. Moreover, extensive and varied datasets are essential to capture diversity in patient demographics, disease distribution, and progression^34–37^.

Federated Learning^38–46^(FL) is a promising solution that enables collaborative model training across multiple sites without sharing sensitive data. In FL, models are trained locally at each site, and their weights are aggregated on a server. This preserves privacy while allowing models to learn from each dataset’s unique information. Despite the theoretical upper bound of performance being incomputable due to privacy constraints, using public datasets allows us to train a central model with centrally aggregated data, setting a benchmark for evaluating FL’s effectiveness. Our approach in this study balances generalizability and optimal site-specific performance. We introduce site-specific fine-tuning FL strategy to enhance the model’s adaptability to both global and local data characteristics. This strategy aims to improve site-specific model generalizability while being comparable to centrally and locally trained model performances, potentially boosting diagnostic capabilities in diverse clinical settings.

## METHODS

This study followed the TRIPOD-AI reporting guidelines as listed in Supplement.

### Datasets

In this paper, we utilized nine public datasets for our FL experiments, comprising Color Fundus Photographs (CFPs) with optic disc and cup segmentations for both glaucoma and non-glaucoma cases. The number of images, country of origin, annotation processes, and citations for each dataset are provided in Supplementary Table 1. The datasets include a total of 5550 CFPs from Bin Rushed^47^ (195 CFP), Chaksu*^48^ (1,345 CFP), Drishti-GS^49^ (101 CFP), G1020^50^ (1,020 CFP), Magrabi*^47^ (94 CFP), Messidor^47^ (460 CFP), ORIGA*^51^ (650 CFP), REFUGE^52^ (1,200 CFP), and Rim-One-DL^53^ (485 CFP), sourced from diverse regions like Saudi Arabia, India, France, Singapore, China, and Spain. Despite incomplete demographic information for these datasets, the inclusion of patient data from seven different countries provides substantial diversity in racial and ethnic representation, imaging specifications, and annotation practices, supporting improved model robustness and generalizability.

Ethical approvals and consent requirements varied across the public datasets. For the Chák□u database, participants were enrolled after obtaining approval from the MAHE Institutional Review Board and Ethics Committee. The study complied with the Declaration of Helsinki, and all subjects (aged 18 to 76 years) provided informed consent. The Messidor, Bin Rushed, and Magrabi datasets were fully de-identified, and research ethics approval for these anonymized data was granted by the University of Waterloo, Canada. For G1020, records were collected retrospectively and anonymized; therefore, patient consent was not required. Similarly, REFUGE images were retrieved from clinical records and anonymized, and the ORIGA dataset was de-identified before deposition. For DRISHTI-GS and RIM-ONE DL, the sources of the datasets do not explicitly report IRB approval or consent status. Each dataset was divided into training, validation, and testing subsets using an 80/10/10 split.

### Algorithmic Development

In this study, we performed a comparative evaluation of three kinds of approaches: (1) local models as the lower bound with no data sharing via site-specific training only, (2) a central model as the upper bound with maximum data aggregation, (3) three standard FL pipelines, against our FL pipeline with site-specific fine-tuning.

### Local models and Central Model

For our upper and lower bound performance, we developed two approaches trained in opposite data sharing conditions: no data sharing and complete data sharing. In detail,

**1. Local models:** We trained a separate model for each site, with each model being trained exclusively on its site’s respective dataset without sharing data between datasets. This strategy had the advantage of returning models tailored specifically to their own dataset.
**2. Central model:** A single model was trained utilizing all datasets combined. This method aggregated the nine publicly available data sources into one single dataset, but at the cost of data-sharing between the sites, which carries security and privacy concerns.

### Federated Learning Models

For training a model within the FL framework, it was assumed that we had multiple sites with local data, each interested in training one model that learns information from its own dataset, with no data leaving each site. FL training kick-started with each site training locally. Once each site had finished the appropriate amount of local training, it sent its local model’s weights to a central server that had the job of aggregating the weights from the various models (e.g., by averaging them, also known as Federated Average^55^) and of sending back the aggregated model to each site. This process, called a “round”, was repeated multiple times. The final result was one overall model trained with no data shared across the sites, but that had learnt from each dataset, nonetheless. This process is depicted in Figure 1a (i) and (ii), where we show the communication between the sites and the server.

**Figure 1:**
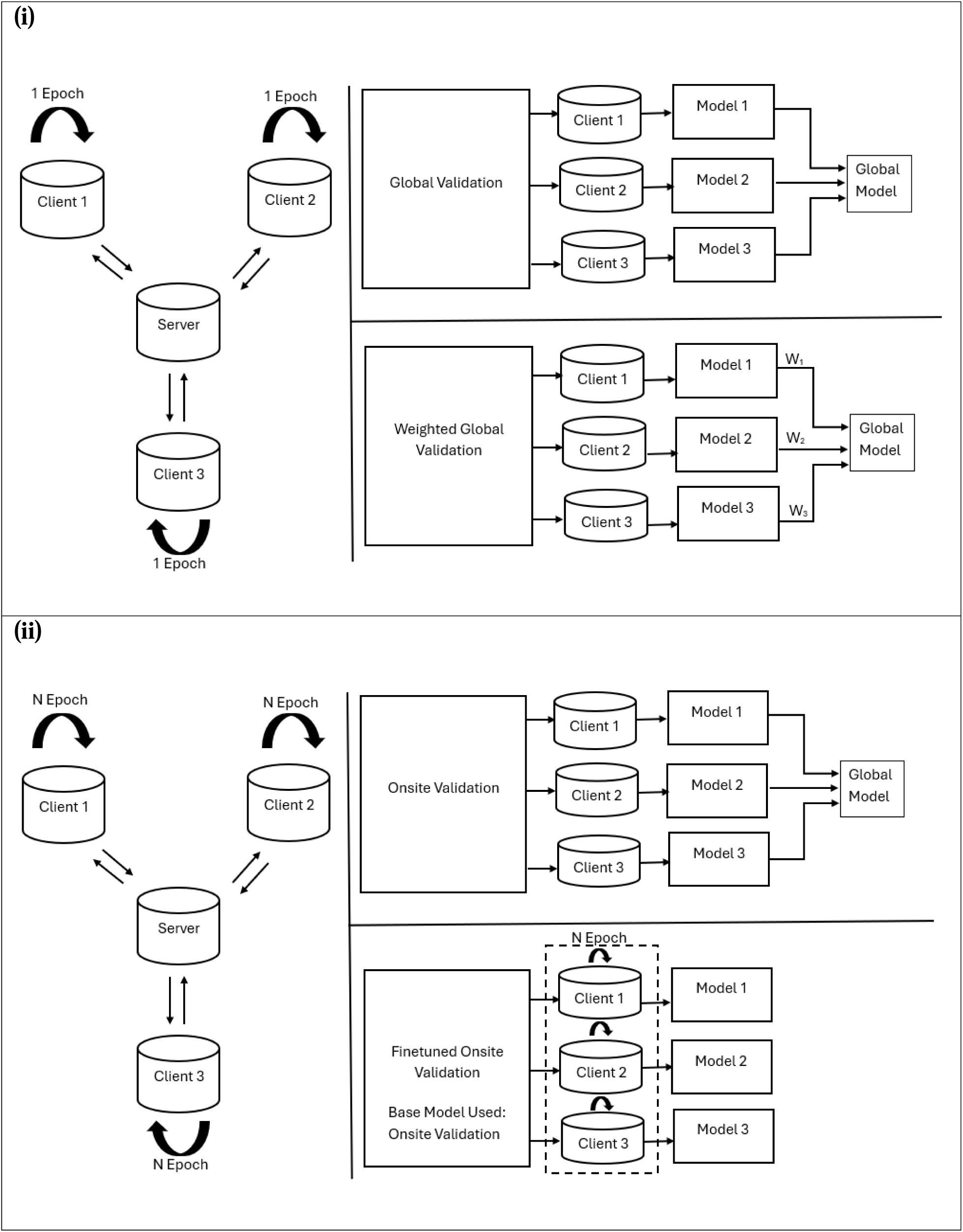
Architectural overview of (i) Global Validation (top right) and Weighted Global Validation (bottom right) (ii) Onsite Validation (top right) and Fine-Tuned Onsite Validation (bottom right) pipelines. In Global Validation, model weights from all clients were averaged, while in Weighted Global Validation, model weights were weighted by site size. In Onsite Validation, each client conducted training and local validation, applying early stopping to select the best model, which was then sent to the server, aggregated, and used for global validation. This Onsite Validation model was used as a starting point to fine-tune on each client’s dataset to generate the nine distinct Fine-tuned Onsite Validation models, optimizing them individually for their respective datasets. a): training phase where each client independently trains local models for 1 (Global Validation / Weighted Global Validation) or multiple epochs (Onsite Validation / Fine-Tuned Onsite Validation). b): Federated Averaging and site-specific fine-tuning phases (for Fine-tuned Onsite Validation only). Models 1, 2, and 3 denote local models for Clients 1, 2, and 3, respectively. For clarity, only three sites (rather than all 9) are represented.

#### 1. FL Pipeline 1: Global Validation

For this FL setup (as shown in Figure 1(i) (b) top), the process involved one epoch of training per site per round, followed by using Federated Averaging (FedAvg) to consolidate all the 9 learnt models into one global model. After each round, the global model was validated on each site, and the validation losses per site were sent to the server, as shown in the top part of Figure 1 (i) (b). The average loss was computed by the server across all sites, and this metric was utilized for early stopping to optimize training (i.e., if the global validation loss did not decrease after a specific number of rounds, the training was interrupted). The advantage of this approach was that it was simple (i.e., 1 epoch of local training per site, evaluation after each round) and no information whatsoever was exchanged regarding each site’s dataset.

#### 2. FL Pipeline 2: Weighted Global Validation

This FL pipeline was built upon the Global Validation setup by introducing a weighted averaging mechanism for model weights (as seen in Figure 1(i) (b), bottom). Here, the weights of models on each site were weighted proportionally to the number of training samples at each site (weights w1, w2, and w3 in Figure 1(i) b bottom); this ensured that the computed average across sites for the model weights reflected the true contribution of each site in terms of the number of samples per site.

#### 3. FL Pipeline 3: Onsite Validation

As shown in Figure 1(ii) (b) top, for this FL setup, each site-specific model was independently trained over several training epochs (instead of 1 epoch). Early stopping was applied to the local training based on the local validation loss to get the best-performing local model. After the local models were sent to the server, the validation loss for the global model was calculated as the mean of the validation losses from these individual sites (similar to the Global Validation approach). This approach was designed with the intuition that choosing the best local models resulted in a better aggregated, central model.

#### 4. FL Pipeline 4: Fine-tuned Onsite Validation

This FL pipeline was built upon the Onsite Validation pipeline by fine-tuning the final Onsite Validation model (i.e., after all training rounds) on each site-specific dataset individually, resulting in nine distinct models (as shown in Figure 1(ii) (b) bottom). The rationale for fine-tuning the Onsite Validation model was that, by fine-tuning on each dataset individually, we adapted the model to site-specific data by tailoring the model’s parameters to better match the unique characteristics of each site’s dataset while also keeping intact the global characteristics from the Onsite Validation model. This strategic adjustment was designed to optimize local accuracy while maintaining robust generalizability (from the federated training before local fine-tuning).

### Model Training Details

The Global Validation model underwent training for 100 rounds, incorporating an early stopping mechanism that halted training if there was no improvement in the global validation loss after 7 rounds. Each round for the Global Validation model involved training the model for one epoch per site. After each epoch, each site’s model’s weights were sent to the central server, where they were averaged. This integrated model was then validated using the global validation approach, and the process was repeated unless early stopping criteria were met.

For the Onsite Validation model, each local model was trained for up to 20 (instead of 1) epochs at its respective site. Early stopping was implemented if there was no improvement in validation metrics after 7 consecutive epochs.

For the Fine-Tuned Onsite Validation models, the Onsite Validation model was fine-tuned on each independent site separately to create site-specific tailored versions of the Onsite Validation model.

### Evaluation and Statistics

We used the Sørensen–Dice coefficient (or Dice score) as our performance metric, which we evaluated on each site’s test set. The global validation dataset used for the validation of the global model was a composite of the test sets from each dataset. When comparing two models, we defined a model to be a winner if the difference in model performance is significant based on the Wilcoxon signed rank test at a significance level of p = 0.5. Bonferroni Correction for multiple hypothesis testing at an alpha value of 0.05 was used for p-value correction.

## RESULTS

### Dataset characteristics

In total, we had 5500 images from a minimum of 917 patients across the 9 datasets. Except for G1020 and RIM-ONE DL, none of the datasets provided details on the number of patients. Demographic characteristics for all 9 datasets, together with the distribution of glaucoma vs healthy, are described in Table 1.

**Table 1:** Demographic characteristics for all public datasets (NS: Not Specified)

| Dataset | # of Patient<br>s | # of Image<br>s | Glaucoma | Healthy | Sex (# of Images) |  | Race/Ethnicity | Age Range |
| --- | --- | --- | --- | --- | --- | --- | --- | --- |
|  |  |  |  |  | Male | Female |  |  |
| Bin Rushed | NS | 195 | NS | NS | 124 | 71 | Saudi-Arabian | 21 to 77 |
| Magrabi | NS | 95 | NS | NS | 47 | 48 | Saudi-Arabian | 9 to 83 |
| MESSIDO<br>R | NS | 460 | NS | NS | NS | NS | NS (collected<br>in France) | NS |
| Chaksu | NS | 1345 | NS | NS | NS | NS | Indian | 18 to<br>76 |
| DRISHTI-<br>GS | NS | 101 | NS | NS | NS | NS | Indian | 40 to<br>80 |
| G1020 | 432 | 1020 | 296 | 724 | NS | NS | NS (collected<br>in Germany) | NS |
| ORIGA-light | NS | 650 | 168 | 482 | NS | NS | Malay | 40 to<br>80 |
| REFUGE | NS | 1200 | 120 | 1080 | NS | NS | Chinese | 21 to<br>77 |
| RIM-ONE<br>DL | 485 | 485 | 172 | 313 | NS | NS | NS (collected<br>in Spain) | NS |

### Model Performance

We conducted a comprehensive comparison of our *Fine-Tuned Onsite Validation FL* pipeline against other approaches (Local models, Central Model, and other standard FL pipelines: Global Validation, Weighted Global Validation, and Onsite Validation) across nine datasets (Figure 2). We also assessed the generalizability of the *Fine-Tuned Onsite Validation* model relative to the corresponding local model when tested on external sites (Figure 3). Detailed performances across individual site-specific local models, the Central Model, and various FL pipelines are provided in Supplementary Tables 2 and 3.

**Figure 2:**
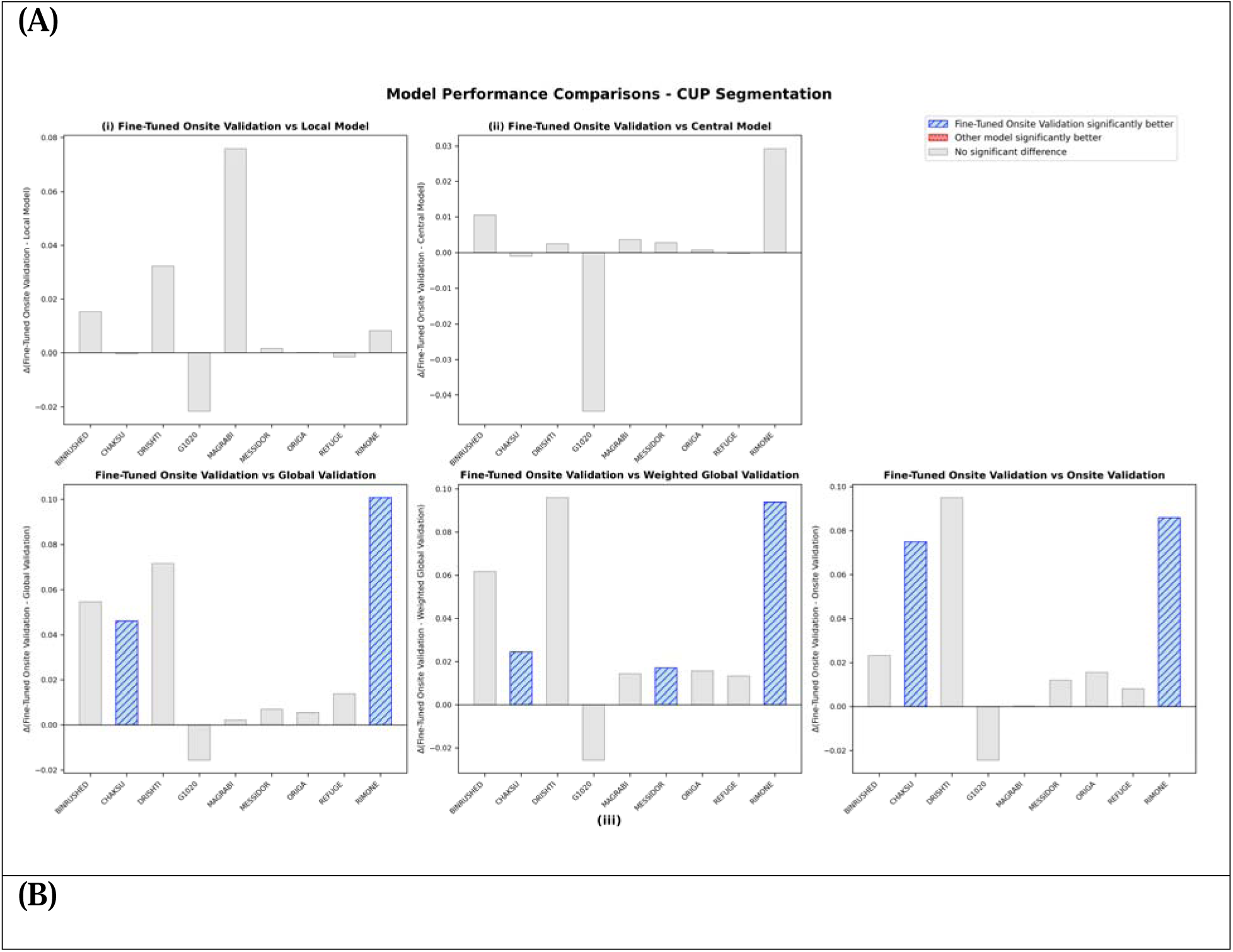
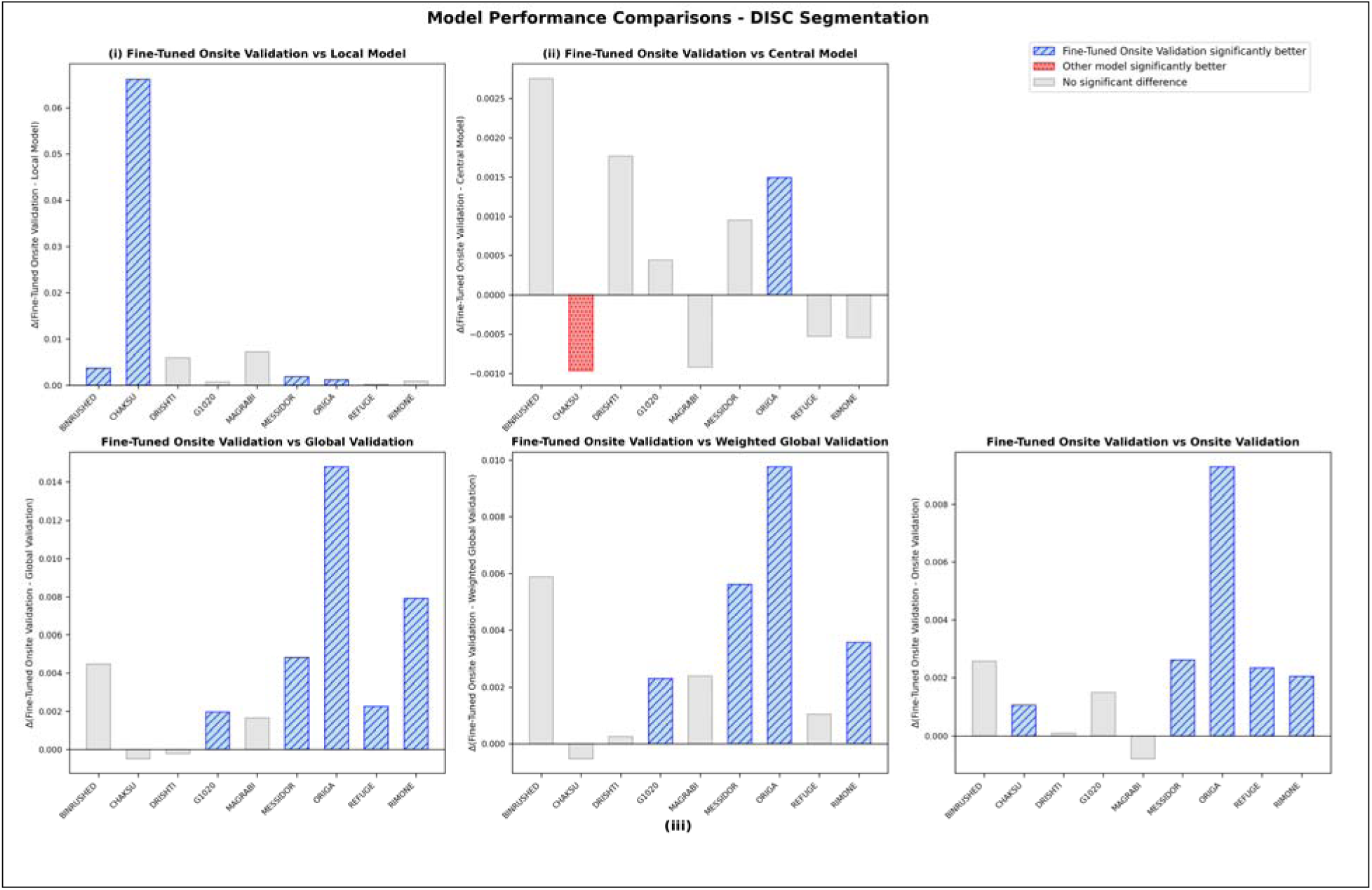
Comparative results for the Fine-Tuned Onsite Validation model vs various approaches. A) Model Comparisons for optic cup segmentation for Fine-Tuned Onsite Validation models when compared to i) local models, ii) the Central model, and iii) standard FL pipelines (Global Validation, Weighted Global Validation, and Onsite Validation). B) Model Comparisons for optic disc segmentation for Fine-Tuned Onsite Validation models when compared to i) local models, ii) Central model, and iii) standard FL pipelines (Global Validation, Weighted Global Validation, and Onsite Validation). For each subplot, X-axis: Delta Difference between mean dice scores of comparator model and Fine-Tuned Onsite Validation model when tested on their respective sites, Y-axis: Datasets used for evaluation of all models and site-specific local models and Fine-Tuned Onsite Validation. The Wilcoxon signed rank test at a significance level of p = 0.05 was used for statistical comparisons of model performances. Bonferroni Correction for multiple hypothesis testing at an alpha value of 0.05 was used for p-value correction. Absolute dice score performances for all models on various sites can be found in supplementary tables 2 and 3.

**Figure 3:**
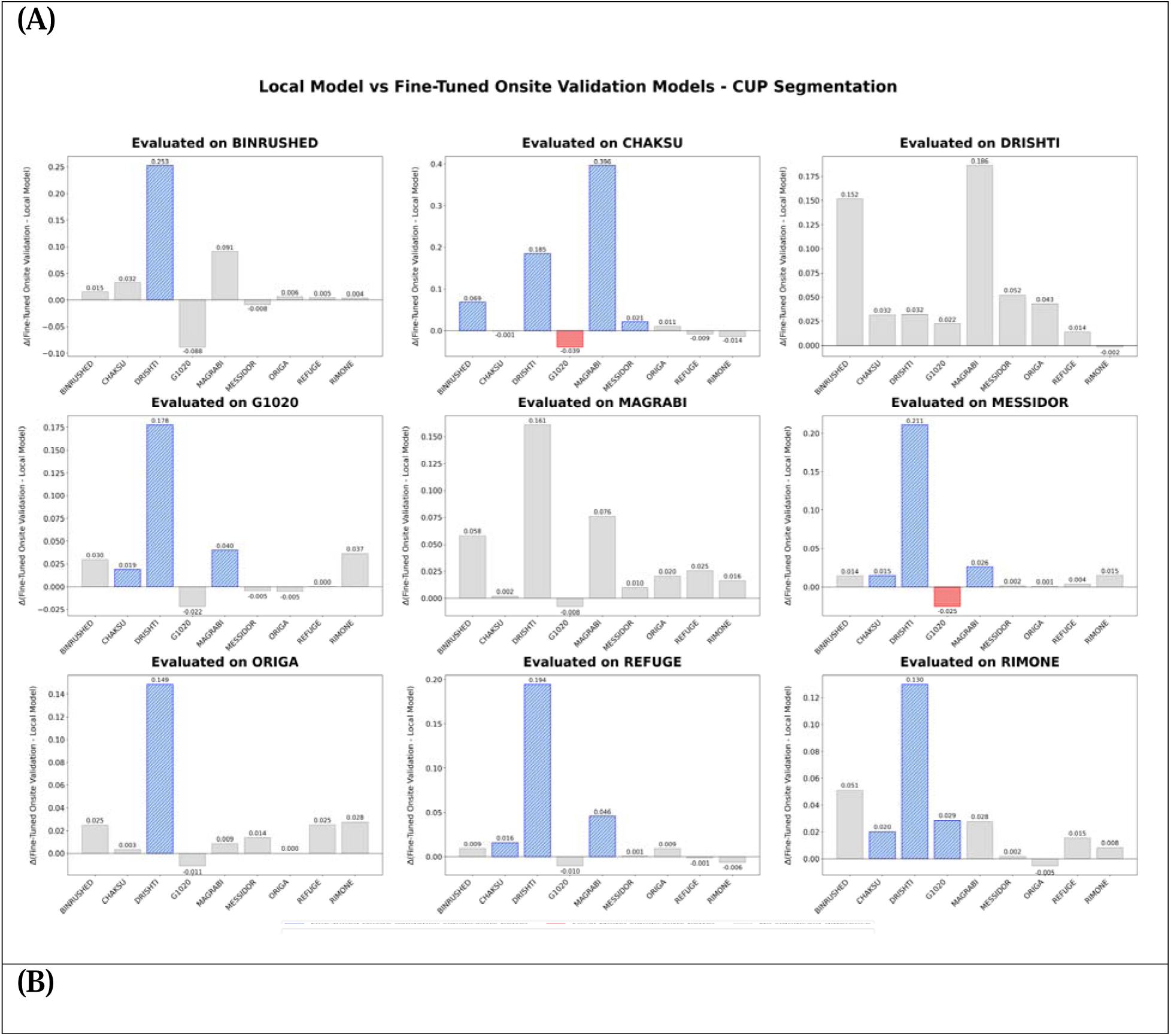
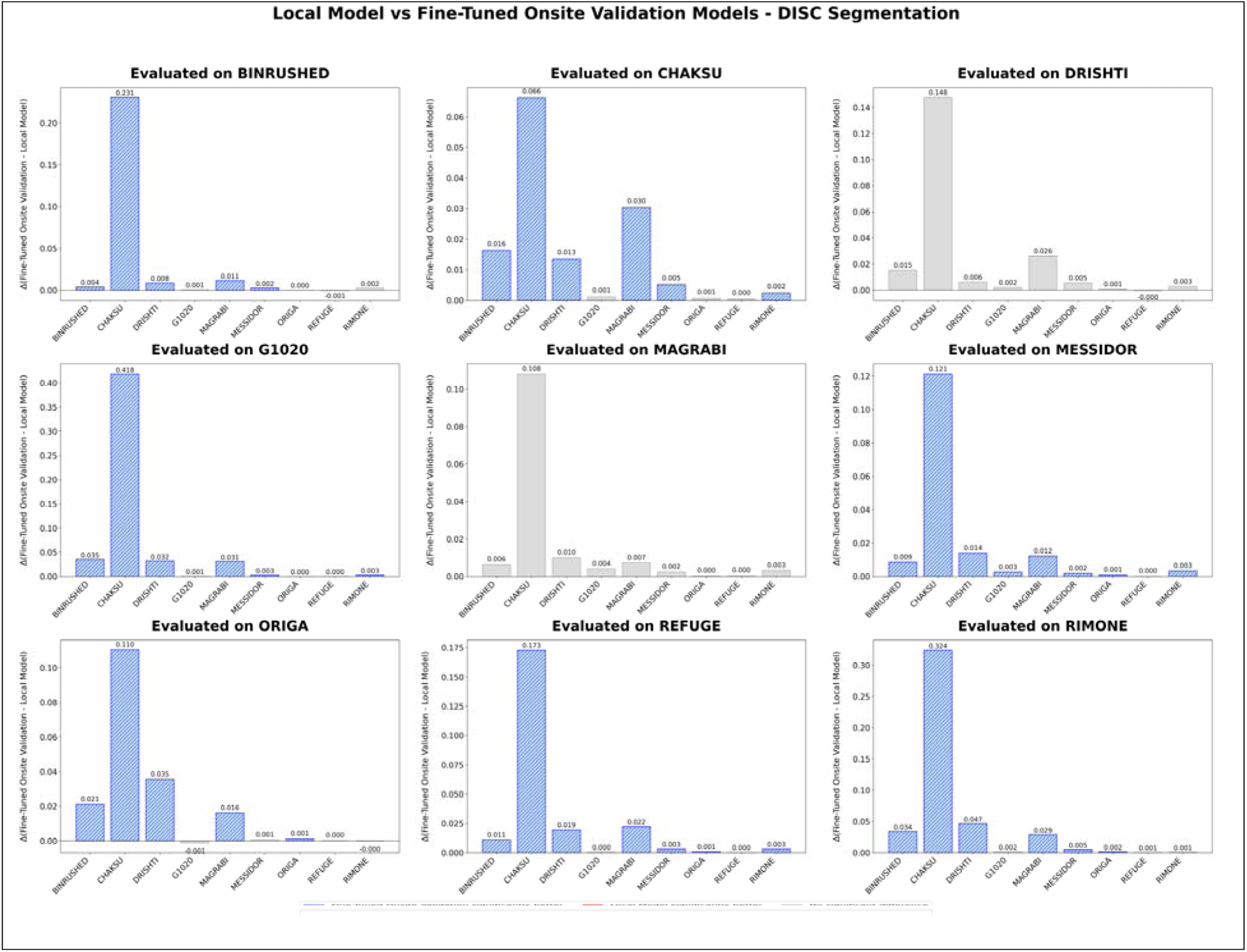
Model generalizability results comparing the Fine-Tuned Onsite Validation model vs the local model when evaluated on external datasets. A) Plots showing Wilcoxon signed-rank test results for optic cup segmentation for Fine-Tuned Onsite Validation models when compared to local models, evaluated on the same dataset as well as external datasets. B) Plots showing Wilcoxon signed-rank test results for optic disc segmentation for Fine-Tuned Onsite Validation models when compared to local models, evaluated on the same dataset as well as external datasets. Each subplot shows an evaluation on a specific dataset. X-axis: Datasets used for site-specific training of local model and Fine-Tuned Onsite Validation FL model, Y-axis: Delta difference between Fine-Tuned Onsite Validation model and local model. The Wilcoxon signed rank test at a significance level of p = 0.05 was used for statistical comparisons of model performances. Bonferroni Correction for multiple hypothesis testing at an alpha value of 0.05 was used for p-value correction. Absolute dice score performances for all models on various sites can be found in supplementary tables 2 and 3.

### Fine-Tuned Onsite Validation Versus Local Models

For cup segmentation (Figure 2A (i)), our *Fine-Tuned Onsite Validation FL model* showed no significant differences against local models across all sites (9/9 ≈ 100%). For disc segmentation (Figure 2B (i)), results were non-significant in 5/9 sites (≈ 55.6%), with the *Fine-Tuned Onsite Validation model* outperforming local models in the remaining 4/9 (≈ 44.4%).

### Fine-Tuned Onsite Validation Versus Central Model

For cup segmentation (Figure 2A (ii)), the *Fine-Tuned Onsite Validation model* did not differ significantly from the Central model at any site (9/9 = 100%). For disc segmentation (Figure 2B (ii)), 7/9 sites (≈ 78%) showed no significant difference; the remaining two sites (ORIGA and CHAKSU) yielded one significant win and one significant loss, respectively.

### Fine-Tuned Onsite Validation Model Versus Standard FL Models

For cup segmentation (Figure 2A (iii)), our *Fine-Tuned Onsite Validation model* achieved significant improvements in 7/27 comparisons (≈ 26%; Global Validation: 2/9, Weighted Global Validation: 3/9, Onsite Validation: 2/9). Gains were concentrated in a few datasets (e.g., CHAKSU, RIMONE, and MESSIDOR against the weighted global variant). For disc segmentation (Figure 2B (iii)), the *Fine-Tuned Onsite Validation model* recorded significant improvements in 14/27 comparisons (≈ 52%; Global Validation: 5/9, Weighted Global Validation: 4/9, Onsite Validation: 5/9), with gains distributed across multiple datasets (G1020, MESSIDOR, ORIGA, REFUGE, RIMONE, etc.).

### Model Generalizability

As shown in Figure 3, we performed 72 pairwise external site comparisons per task, evaluating each training site’s *Fine-Tuned Onsite Validation* model against the site-specific local model on the same external dataset. We reported: a) Optic cup (Figure 3A): Significant wins in 25.0% (18/72), losses in 2.8% (2/72), and no significant difference in 72.2% (52/72). b) Optic disc (Figure 3B): Significant wins in 54.2% (39/72), no losses, and no significant difference in 45.8% (33/72). Dataset-level disc improvements were most pronounced for ORIGA (87.5% wins), REFUGE (87.5%), and MESSIDOR (75%), with no significant changes for BINRUSHED or CHAKSU. For the cup, gains were smaller and varied across datasets (e.g., REFUGE: 50% wins, MESSIDOR: 37.5% wins, no changes for BINRUSHED or CHAKSU).

## DISCUSSION

In this work, we demonstrated how an FL-based site-specific fine-tuning model for optic disc and cup segmentation from color fundus photographs not just outperforms other standard FL pipelines but also achieves performance comparable to the central model, while preserving onsite data privacy and demonstrating enhanced generalizability relative to site-specific, local models.

The Central model, trained on pooled multi-site data, provides a robust benchmark for FL pipelines, as it more effectively accounts for dataset heterogeneity than local models trained at individual sites. Our findings indicate that our *Fine-tuned Onsite validation* FL models achieve performance comparable to the Central model on the respective local test sets of each site (Figure 2(ii)). These results align with recent studies^59–61^ demonstrating that locally adapted FL models can closely match, and in some cases exceed, centralized approaches on specific performance metrics. Such improvements are largely attributable to their capacity to accommodate site-specific data variations without necessitating direct data sharing, thereby preserving patient privacy and ensuring compliance with regulatory requirements.

Compared with site-specific local models, our *Fine-tuned Onsite Validation* FL model showed no significant difference across the majority of the sites (Figure 2(i)). While local models perform competitively in optic cup segmentation (likely because local models already capture site-specific traits, leaving limited scope for the *Fine-Tuned Onsite Validation*), this phenomenon does not extend to disc segmentation. In this setting, initializing from a globally trained FL model and applying site-specific fine-tuning outperforms training from scratch on isolated data. This is consistent with prior studies^62,63^ showing that FL global models provide robust, generalizable feature representations that serve as stronger starting points for local adaptation. Although standard FL pipelines (Global Validation, Weighted Global Validation, and Onsite Validation) yield strong overall results, they often underfit site-specific heterogeneity. By contrast, our *Fine-tuned Onsite Validation FL model* introduces a critical personalization step, improving upon the strongest baseline (Onsite Validation), particularly in optic disc segmentation tasks (Figure 2(iii)).

In terms of external site generalizability, local models (Supplementary Tables 2–3) performed well within their site-specific distributions but showed limited cross-site generalizability. This limitation was most pronounced in optic cup segmentation (Supplementary Table 3), where in-domain performance (diagonal entries) substantially exceeded out-of-domain performance (off-diagonal entries). In contrast, our *Fine-tuned Onsite Validation* FL model consistently outperformed site-specific local models in disc segmentation external evaluations (Figure 3B), achieving significant wins in over half of all comparisons without performance degradation. For cup segmentation (Figure 3A), improvements were more modest, as local models were already near-optimal, limiting the benefits of the *Fine-Tuned Onsite Validation FL model* with small site-specific datasets. This discrepancy likely reflects underlying biological and imaging factors: optic cup segmentation is inherently more challenging due to vessel density, gradual intensity transitions at the cup-rim boundary, greater inter-rater variability, and glaucomatous changes in cup morphology^64^. Consequently, the cup is more susceptible than the disc to inter-site heterogeneity, reducing the utility of meaningful generalized representations learned from pooled or federated sources.

FL has transformed privacy-preserving machine learning in multi-institutional medical settings^62,65–68^. However, it can exhibit bias toward larger datasets, which may reduce performance on smaller sites^29,40,69,70^. This issue is especially relevant in glaucoma detection, where heterogeneity in disease presentation and diagnostic workflows across sites can hinder both training and generalization^71–75^. Our study advances FL by incorporating site-specific fine-tuning, differentiating our approach from traditional methods. This site-specific fine-tuning of FL consistently matched the performance of the Central Model while outperforming all other standard FL pipelines and site-specific local models across diverse datasets. It preserved performance for cup segmentation and delivered substantial gains for disc segmentation, both in within-site evaluations and in cross-site generalizability tests, achieving significant wins in over half of all external comparisons without any losses.

### Future Work/Limitations

We tested our methods on publicly available data; doing so allows us to compute an upper bound for our federated approaches (by training a central model). However, reliance on these public datasets is constrained by incomplete demographic information, which limits the ability to evaluate fairness or identify potential biases across subpopulations in the absence of detailed racial and disease distribution data. Future work includes validating our FL approaches in real-world clinical settings to better understand their performance in less controlled environments compared to curated public datasets. At the same time, our preprocessing requires a detection model to identify the ONH; future experiments will explore the elimination of this intermediate step. Additionally, cropping around the ONH may be excluding important and useful diagnostic information that could be used for glaucoma evaluation. Future work could additionally include other imaging modalities like OCTs for diagnosis and monitoring. And lastly, future work could also explore the variability in cup vs disc segmentation performance.

## CONCLUSION

Our findings demonstrate that federated learning with site-specific fine-tuning can achieve central-level performance while preserving data privacy and can substantially improve cross-site generalizability. This approach enables the development of robust, adaptable segmentation models that leverage diverse, multi-institutional datasets without data sharing, supporting standardized and scalable glaucoma detection and monitoring across heterogeneous clinical settings.

## Data Availability

All the code for the pipelines used can be found here:
https://github.com/QTIM-Lab/federatedLearning_glaucoma_segmentation/tree/main

## DATA SHARING STATEMENT

All the code for the pipelines used can be found here: https://github.com/QTIM-Lab/federatedLearning_glaucoma_segmentation/tree/main

## Access to data and data analysis

Mr. Shrivastava and Dr. Singh had full access to all the data in the study and take responsibility for the integrity of the data and the accuracy of the data analysis.

