## Supplementary for "A Federated Learning-based Optic Disc and Cup Segmentation Model for Glaucoma Monitoring In Color Fundus Photographs"

**SUPPLEMENTARY MATERIAL**

**Weighted Global Validation**

Mathematically, this methodology is encapsulated as follows:

Let S be the set of all sites, 𝑁 represent the total number of samples across all sites, and 𝑛𝑖 represent the number of training samples at site 𝑖. For each site 𝑖 ∈ 𝑆, during the calculation of average model weights, each site model $weight_{i}$ is normalized by:

$weight$ = (𝑛𝑖/𝑁) * $weight_{i}$

where:

- $weight_{i}$ is the original site-specific model weights for site i,
- 𝑁 is the total number of training samples for all sites,
- 𝑛𝑖 is the number of training samples at site i, and
- $weight$is the averaged global model weights.

### **Image Preprocessing**

We preprocess each dataset to ensure consistency in image quality and size in order to reduce model bias and improve the generalizability of our findings, especially in a Federated Learning setup (where images could come from multiple sources and have different formats and standards). The preprocessing steps include:

1. Resizing images to a uniform size of 487x487 pixels.
2. Centering on the Optic Nerve Head (ONH) using a YOLOv8^57^ object detector trained on the combined nine datasets, which follows the same 80/10/10 split for training, validation, and test sets as our FL experiments. This centers crucial anatomical features for glaucoma analysis.
3. Adding padding to standardize dimensions to 512x512 pixels, accommodating any variations in ONH placement across images and ensuring uniform input for segmentation models.
4. For datasets with multiple expert annotations, the STAPLE54 method, an Expectation-Maximization algorithm, was used to aggregate consensus segmentation labels to enhance accuracy and consistency.

### **Model architecture and Loss function**

Segmentation models in all categories used an ade22k pretrained Mask2Former (Swin Transformer backbone) model^55,56^ with a multi-class loss function (categories: background, cup, and disc) and AdamW^57^ optimizer. This Mask2Former model was pre-trained on the dataset Cityscapes for semantic segmentation^58^.

**Table 1: Dataset details (NS: Not Specified)**

| Dataset | Image Acquisition Dates | Acquisition Location | Imaging Devices & Parameters | Study Methodology and Constraints | Source | Annotators |
| --- | --- | --- | --- | --- | --- | --- |
| Bin  Rushed | 2014 | Bin Rushed Ophthalmic Center in Riyadh, Saudi Arabia | Canon CR2 non-mydriatic digital retinal camera; Images are 2376X1584p | The dataset includes randomly chosen normal and glaucomatous fundus images | [https://commons. datacite.org/doi.o rg/10.7302/Z23R](https://commons.datacite.org/doi.org/10.7302/Z23R0R29)  [0R29](https://commons.datacite.org/doi.org/10.7302/Z23R0R29),  [https://deepblue.l ib.umich.edu/dat a/concern/data_s ets/3b591905z](https://deepblue.lib.umich.edu/data/concern/data_sets/3b591905z) | 6 experienced ophthalmologists. These six annotators were specialists from different eye centers in Saudi Arabia |
| Chaksu | NS | Department of Ophthalmology at Kasturba Medical College (KMC), Manipal, India, and other institutions in Manipal, Karnataka, India | Acquired using three brands of commercially available non-mydriatic fundus cameras: Remidio (2448 × 3264 pixels), Forus (2048 × 1536 pixels), and Bosch (1920 × 1440 pixels). Images have an approximate 40-degree Field of View (FoV). | The fundus examination was conducted undilated. Images are approximately Optic Disc (OD)-centered. | [https://www.ncbi. nlm.nih.gov/pmc/ articles/PMC989](https://www.ncbi.nlm.nih.gov/pmc/articles/PMC9898274/)  [8274/](https://www.ncbi.nlm.nih.gov/pmc/articles/PMC9898274/) | 5 expert Indian ophthalmologists. Specifically, the annotator team included: 2 experienced Professors, 2 Associate Professors and 1 clinical practitioner. 3 of these experts were glaucoma specialists, and 2 were general ophthalmologists. Their clinical experience ranged from 5 to 15 years. |
| Drishti-GS | NS | Aravind Eye Hospital, Madurai, India | Camera: NS; Images are 2896 × 1944 pixels. The collection utilized a 30-degree field-of-view. | Images were taken with eyes dilated. The images were centered on the OD. Images with poor contrast, positioning of OD/OC too eccentric, or containing certain retinal artifacts were discarded. | [https://www.kagg le.com/datasets/l okeshsaipureddi/ drishtigs-retina-d ataset-for-onh-se gmentation](https://www.kaggle.com/datasets/lokeshsaipureddi/drishtigs-retina-dataset-for-onh-segmentation), [https://ieeexplore .ieee.org/docum ent/6867807](https://ieeexplore.ieee.org/document/6867807) | 4 different human experts. These experts possessed varying clinical experience of 3, 5, 9, and 20 years. |
| . G1020 | 2005-2017 | Private clinical practice in Kaiserslautern, Germany | Camera: NS; Images were taken with a 45-degree field of view. Images were stored in JPG format, with final fundus regions sized between 1944×2108 and 2426×3007 pixels | Images were collected retrospectively and taken after using dilation drops. No specific imaging constraints were imposed (e.g., centering the OD or macula), reflecting routine clinical practice. | [https://arxiv.org/a bs/2006.09158](https://arxiv.org/abs/2006.09158) | The segmentation ground truth was initially marked by an unnamed expert. These manual annotations were then verified and corrected (if necessary) by a veteran ophthalmologist with more than 25 years of clinical experience. |
| Magrabi | 2012-2014 | Magrabi Eye center, Riyadh, Saudi Arabia | Topcon TRC 50DX mydriatic retinal camera. Images are 2743X1936p. | Both glaucomatous and normal healthy images were chosen randomly | [https://commons. datacite.org/doi.o](https://commons.datacite.org/doi.org/10.7302/Z23R0R29)  [rg/10.7302/Z23R](https://commons.datacite.org/doi.org/10.7302/Z23R0R29)  [0R29](https://commons.datacite.org/doi.org/10.7302/Z23R0R29) | 6 experienced ophthalmologists who were specialists from different eye centers in Saudi Arabia. |
| Messidor | 2004 | France | The images have variable resolutions (e.g., 2304 × 1536, 2240 × 1488, 1440 × 960 pixels). The FOV is 45 degrees. | The total MESSIDOR database (1200 images) included 800 images acquired with pupil dilation and 400 without dilation. | [https://commons. datacite.org/doi.o](https://commons.datacite.org/doi.org/10.7302/Z23R0R29)  [rg/10.7302/Z23R](https://commons.datacite.org/doi.org/10.7302/Z23R0R29)  [0R29](https://commons.datacite.org/doi.org/10.7302/Z23R0R29) | single clinician. |
| ORIGA | 2004-2007 | Collected as part of the Singapore Malay Eye Study (SIMES), Singapore | Camera: NS | The images were collected in a population-based study. The dataset includes images labeled for glaucoma severity grading. | [https://www.kagg le.com/datasets/ arnavjain1/glauc oma-datasets](https://www.kaggle.com/datasets/arnavjain1/glaucoma-datasets) | Trained professionals from Singapore Eye Research Institute. |
| REFUGE | NS | Retrieved retrospectively from multiple sources including several hospitals and clinical studies in China | Two devices were used: a Zeiss Visucam 500 camera (2124 × 2056 pixels) and a Canon CR-2 device (1634 × 1634 pixels). The FOV is 45 degrees. | Only high-quality images were selected. Images are centered at the posterior pole, ensuring both the macula and optic disc are visible. The non-glaucomatous set explicitly includes cases with comorbidities like diabetic retinopathy, myopia, and megalopapilae. Glaucoma labels were assigned based on a comprehensive evaluation of clinical records (including follow-up images, IOP, OCT, and visual fields), not just the single fundus photograph. | [https://ieee-data port.org/docume nts/refuge-retinal -fundus-glaucom a-challenge](https://ieee-dataport.org/documents/refuge-retinal-fundus-glaucoma-challenge) | 7 independent glaucoma specialists from the Zhongshan Ophthalmic Center (Sun Yat-sen University, China). Their average experience was 8 years, ranging from 5 to 10 years. A senior specialist with more than 10 years of experience in glaucoma performed a quality check of the resulting segmentations. |
| Rim-One-DL | 2015 for part of the dataset (RIM-One v3) | Collected at three Spanish hospitals: Hospital Universitario de Canarias (HUC), Hospital Universitario Miguel Servet (HUMS), and Hospital Clínico Universitario San Carlos (HCSC) | Canon EOS 40D Mark II, Canon WX-D stereo fundus camera. The FOV is 45 degrees. | Images from RIM-One v3 were acquired with a horizontal FOV of 20° and a vertical FOV of 27°, focused on the ONH area. The final DL dataset was carefully curated by removing duplicates and ensuring only one image per patient from v3 was used. | [https://medimrg.webs.ull.es/](https://medimrg.webs.ull.es/%22%20/t%20%22_blank), [https://www.ias-iss.org/ojs/IAS/article/view/2346](https://www.ias-iss.org/ojs/IAS/article/view/2346%22%20/t%20%22_blank) | RIM-ONE v1: 5 experts in the field.  RIM-ONE v2: 2 medical specialist.  RIM-ONE DL: 2 experts reviewed all the images, followed by a discussion with a 3^rd^ specialist to make the final decision. |

**Table 2: Results for optic disc segmentation. Table shows results for locally trained models b) Table shows results for FL fine-tuned models (on onsite validation FL model)** [The datasets 1-9 are in this order: Bin Rushed, Chaksu, Drishti, G1020, Magrabi, Messidor, Origa, Refuge, and Rim-One]

**a)**

|  |  | **Testing →**  (test) | | | | | | | | |
| --- | --- | --- | --- | --- | --- | --- | --- | --- | --- | --- |
|  | **Datasets/Sites** | **1**  **(20)** | **2**  **(135)** | **3**  **(11)** | **4 (102)** | **5**  **(10)** | **6**  **(46)** | **7**  **(65)** | **8 (120)** | **9**  **(49)** |
| **Training**  **(train, val) ↓** | **1 (156, 19)** | 0.9819 | 0.9582 | 0.9655 | 0.9431 | 0.9795 | 0.9769 | 0.9632 | 0.9670 | 0.9491 |
|  | **2**  **(1076, 134)** | 0.7515 | 0.9177 | 0.8400 | 0.5592 | 0.8746 | 0.8601 | 0.8710 | 0.8111 | 0.6554 |
|  | **3 (80,10)** | 0.9728 | 0.9692 | 0.9811 | 0.9465 | 0.9740 | 0.9683 | 0.9504 | 0.9649 | 0.9374 |
|  | **4**  **(816, 102)** | 0.9829 | 0.9801 | 0.9828 | 0.9796 | 0.9824 | 0.9815 | 0.9838 | 0.9816 | 0.9796 |
|  | **5 (75, 9)** | 0.9733 | 0.9445 | 0.9532 | 0.9463 | 0.9774 | 0.9722 | 0.9657 | 0.9568 | 0.9538 |
|  | **6 (368, 46)** | 0.9846 | 0.9684 | 0.9741 | 0.9757 | 0.9838 | 0.9842 | 0.9830 | 0.9754 | 0.9768 |
|  | **7 (520, 65)** | 0.9746 | 0.9806 | 0.9823 | 0.9727 | 0.9788 | 0.9756 | 0.9949 | 0.9858 | 0.9848 |
|  | **8**  **(960, 120)** | 0.9794 | 0.9820 | 0.9849 | 0.9746 | 0.9806 | 0.9793 | 0.9921 | 0.9875 | 0.9854 |
|  | **9(388, 48)** | 0.9799 | 0.9764 | 0.9819 | 0.9742 | 0.9819 | 0.9807 | 0.9880 | 0.9788 | 0.9852 |
|  | **Central model** | 0.9829 | 0.9849 | 0.9854 | 0.9799 | 0.9855 | 0.9850 | 0.9947 | 0.9882 | 0.9867 |
|  | **Global Validation** | 0.9812 | 0.9844 | 0.9873 | 0.9784 | 0.9829 | 0.9812 | 0.9813 | 0.9854 | 0.9782 |
|  | **Weighted Global Validation** | 0.9798 | 0.9845 | 0.9869 | 0.9780 | 0.9822 | 0.9804 | 0.9864 | 0.9867 | 0.9825 |
|  | **Onsite Validation** | 0.9831 | 0.9829 | 0.9870 | 0.9788 | 0.9854 | 0.9834 | 0.9869 | 0.9854 | 0.9841 |

**b)**

|  |  | **Testing →**  (test) | | | | | | | | |
| --- | --- | --- | --- | --- | --- | --- | --- | --- | --- | --- |
|  | **Datasets/Sites** | **1**  **(20)** | **2 (135)** | **3**  **(11)** | **4 (102)** | **5**  **(10)** | **6**  **(46)** | **7**  **(65)** | **8 (120)** | **9**  **(49)** |
| **Training**  **(train, val) ↓** | **1 (156, 19)** | 0.9857 | 0.9745 | 0.9807 | 0.9783 | 0.9859 | 0.9854 | 0.9843 | 0.9780 | 0.9830 |
|  | **2**  **(1076, 134)** | 0.9821 | 0.9840 | 0.9877 | 0.9775 | 0.9826 | 0.9813 | 0.9814 | 0.9839 | 0.9793 |
|  | **3 (80,10)** | 0.9812 | 0.9827 | 0.9871 | 0.9785 | 0.9840 | 0.9823 | 0.9859 | 0.9844 | 0.9839 |
|  | **4**  **(816, 102)** | 0.9835 | 0.9812 | 0.9846 | 0.9803 | 0.9864 | 0.9840 | 0.9826 | 0.9819 | 0.9813 |
|  | **5 (75, 9)** | 0.9846 | 0.9748 | 0.9793 | 0.9769 | 0.9846 | 0.9844 | 0.9818 | 0.9788 | 0.9826 |
|  | **6 (368, 46)** | 0.9868 | 0.9735 | 0.9796 | 0.9786 | 0.9862 | 0.9860 | 0.9835 | 0.9784 | 0.9816 |
|  | **7 (520, 65)** | 0.9747 | 0.9811 | 0.9830 | 0.9730 | 0.9791 | 0.9765 | 0.9962 | 0.9867 | 0.9863 |
|  | **8**  **(960, 120)** | 0.9789 | 0.9822 | 0.9845 | 0.9751 | 0.9811 | 0.9794 | 0.9923 | 0.9877 | 0.9859 |
|  | **9 (388, 48)** | 0.9821 | 0.9787 | 0.9846 | 0.9774 | 0.9851 | 0.9840 | 0.9878 | 0.9818 | 0.9861 |

**Table 3: Results for optic cup segmentation a) Table showing results for locally trained model b) Table showing results for FL fine-tuned models (on onsite validation FL model)** [The datasets 1-9 are in this order: Bin Rushed, Chaksu, Drishti, G1020, Magrabi, Messidor, Origa, Refuge, and Rim-One]

**a)**

|  |  | **Testing →**  (test) | | | | | | | | |
| --- | --- | --- | --- | --- | --- | --- | --- | --- | --- | --- |
|  | **Datasets/Sites** | **1**  **(20)** | **2 (135)** | **3**  **(11)** | **4 (102)** | **5**  **(10)** | **6**  **(46)** | **7**  **(65)** | **8 (120)** | **9**  **(49)** |
| **Training**  **(train, val)**  **↓** | **1 (156, 19)** | 0.8947 | 0.6822 | 0.6368 | 0.6030 | 0.8521 | 0.8885 | 0.8144 | 0.8667 | 0.7046 |
|  | **2**  **(1076, 134)** | 0.7515 | 0.9177 | 0.8400 | 0.5592 | 0.8746 | 0.8601 | 0.8710 | 0.8111 | 0.6554 |
|  | **3 (80,10)** | 0.5782 | 0.7119 | 0.9100 | 0.4069 | 0.7264 | 0.6759 | 0.7174 | 0.6350 | 0.5461 |
|  | **4**  **(816, 102)** | 0.8745 | 0.7353 | 0.7370 | 0.6274 | 0.8897 | 0.8649 | 0.8010 | 0.8663 | 0.7178 |
|  | **5 (75, 9)** | 0.8052 | 0.4254 | 0.6470 | 0.5847 | 0.8543 | 0.8928 | 0.8550 | 0.8357 | 0.7114 |
|  | **6 (368, 46)** | 0.8864 | 0.8020 | 0.7490 | 0.6235 | 0.9125 | 0.9275 | 0.8512 | 0.8777 | 0.7262 |
|  | **7 (520, 65)** | 0.8151 | 0.8815 | 0.8567 | 0.5913 | 0.8854 | 0.8925 | 0.8845 | 0.8384 | 0.6914 |
|  | **8**  **(960, 120)** | 0.8774 | 0.8282 | 0.8408 | 0.6328 | 0.8900 | 0.8811 | 0.8217 | 0.8958 | 0.7227 |
|  | **9** | 0.7687 | 0.6799 | 0.8155 | 0.5873 | 0.8207 | 0.7856 | 0.7567 | 0.8168 | 0.8150 |
|  | **(388, 48)** |  |  |  |  |  |  |  |  |  |
|  | **Central model** | 0.8994 | 0.9182 | 0.9396 | 0.6503 | 0.9266 | 0.9263 | 0.8838 | 0.8947 | 0.7941 |
|  | **Global Validation** | 0.8555 | 0.8711 | 0.8706 | 0.6213 | 0.9281 | 0.9221 | 0.8790 | 0.8804 | 0.7224 |
|  | **Weighted Global Validation** | 0.8482 | 0.8927 | 0.8462 | 0.6315 | 0.9160 | 0.9121 | 0.8689 | 0.8811 | 0.7295 |
|  | **Onsite Validation** | 0.8866 | 0.8423 | 0.8471 | 0.6302 | 0.9301 | 0.9172 | 0.8691 | 0.8866 | 0.7376 |

**b)**

|  |  | **Testing →**  (test) | | | | | | | | |
| --- | --- | --- | --- | --- | --- | --- | --- | --- | --- | --- |
|  | **Datasets/Sites** | **1**  **(20)** | **2 (135)** | **3**  **(11)** | **4 (102)** | **5**  **(10)** | **6**  **(46)** | **7**  **(65)** | **8 (120)** | **9**  **(49)** |
| **Training**  **(train, val)**  **↓** | **1 (156, 19)** | 0.9100 | 0.7508 | 0.7884 | 0.6327 | 0.9102 | 0.9030 | 0.8392 | 0.8761 | 0.7557 |
|  | **2**  **(1076, 134)** | 0.7838 | 0.9172 | 0.8715 | 0.5781 | 0.8764 | 0.8749 | 0.8742 | 0.8270 | 0.6755 |
|  | **3 (80,10)** | 0.8307 | 0.8971 | 0.9421 | 0.5845 | 0.8873 | 0.8868 | 0.8659 | 0.8294 | 0.6759 |
|  | **4**  **(816, 102)** | 0.7863 | 0.6962 | 0.7591 | 0.6057 | 0.8820 | 0.8395 | 0.7900 | 0.8560 | 0.7464 |
|  | **5 (75, 9)** | 0.8962 | 0.8216 | 0.8332 | 0.6248 | 0.9303 | 0.9189 | 0.8635 | 0.8815 | 0.7392 |
|  | **6 (368, 46)** | 0.8782 | 0.8233 | 0.8012 | 0.6189 | 0.9223 | 0.9291 | 0.8650 | 0.8784 | 0.7280 |
|  | **7 (520, 65)** | 0.8215 | 0.8927 | 0.8997 | 0.5860 | 0.9059 | 0.8936 | 0.8845 | 0.8476 | 0.6862 |
|  | **8**  **(960, 120)** | 0.8821 | 0.8196 | 0.8546 | 0.6330 | 0.9155 | 0.8848 | 0.8467 | 0.8944 | 0.7381 |
|  | **9 (388, 48)** | 0.7727 | 0.6661 | 0.8138 | 0.6239 | 0.8368 | 0.8007 | 0.7842 | 0.8107 | 0.8233 |

**Figure 1: Comparative performance results for standard FL models vs central and local models.**

(a) **Bar plots showing difference of mean dice scores and Wilcoxon signed-rank test significant winners for optic disc segmentation for standard FL models when compared to 1) local model and 2) central model evaluated on the same dataset.**

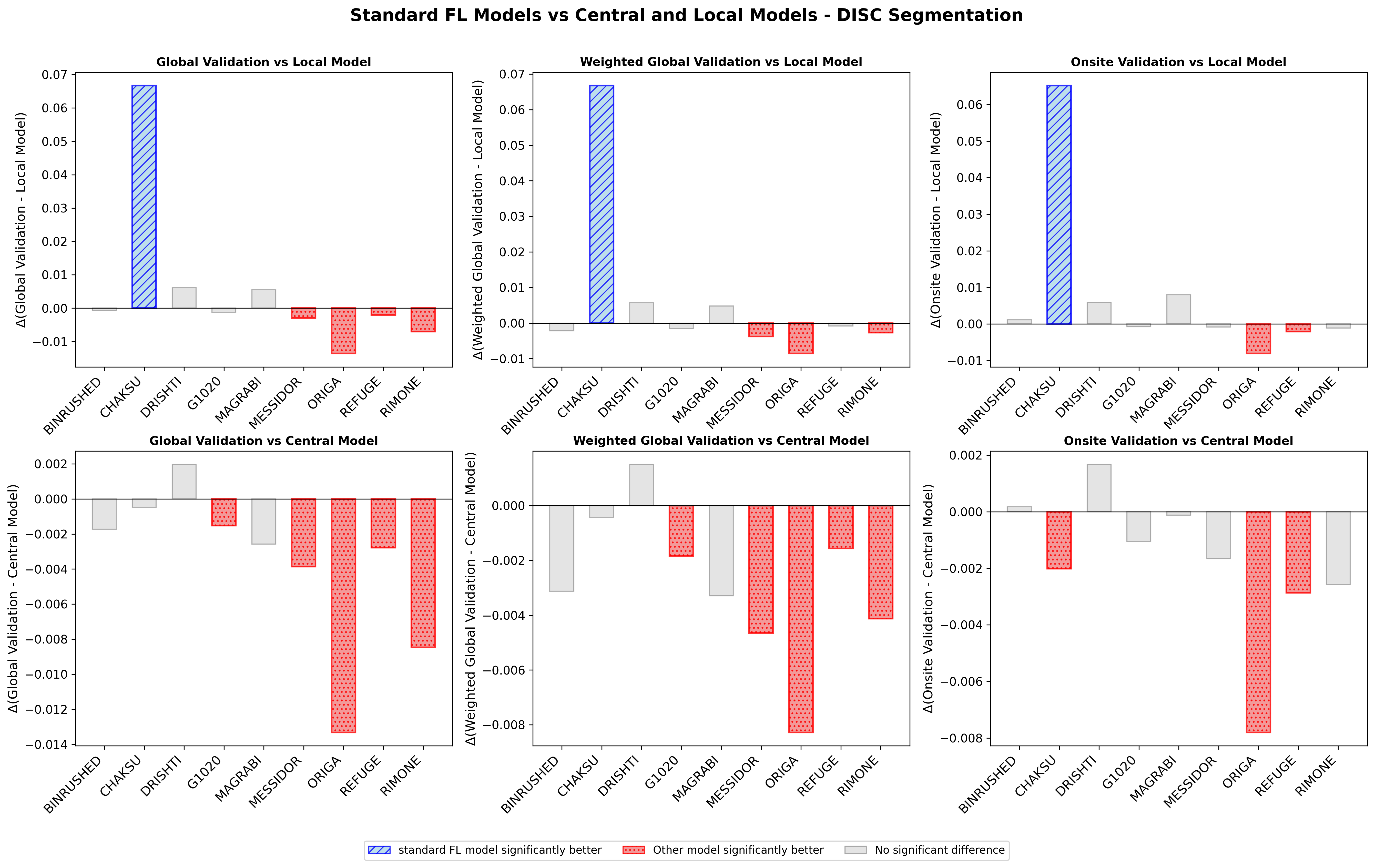

(b) **Bar plots showing difference of mean dice scores and Wilcoxon signed-rank test significant winners for optic cup segmentation for standard FL models when compared to 1) local modeland 2) central model evaluated on the same dataset.**

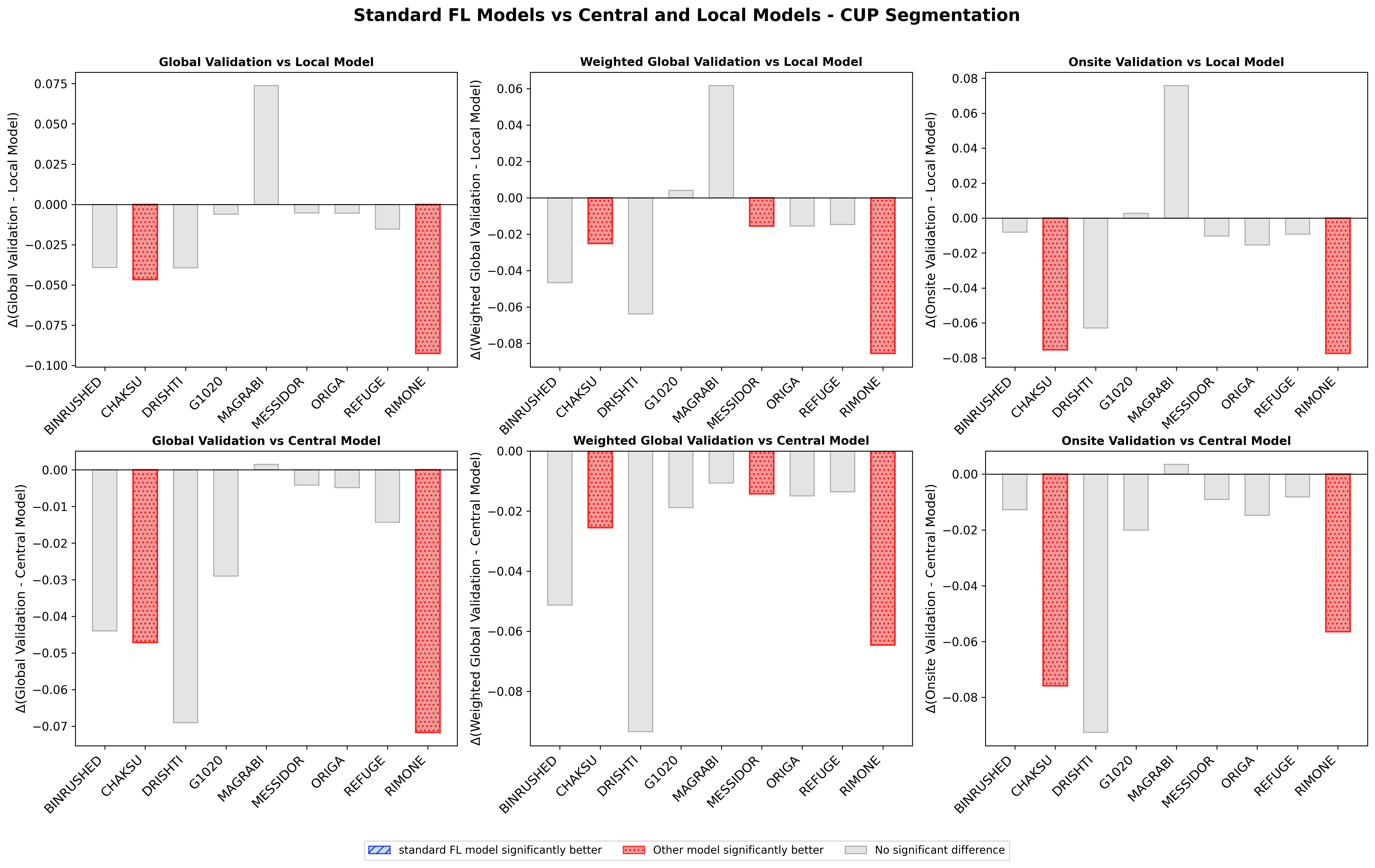

**Figure 2: Comparative performance results for Central Model vs Localmodel evaluated on the same dataset for (1) Cup segmentation and (2) Disc segmentation**

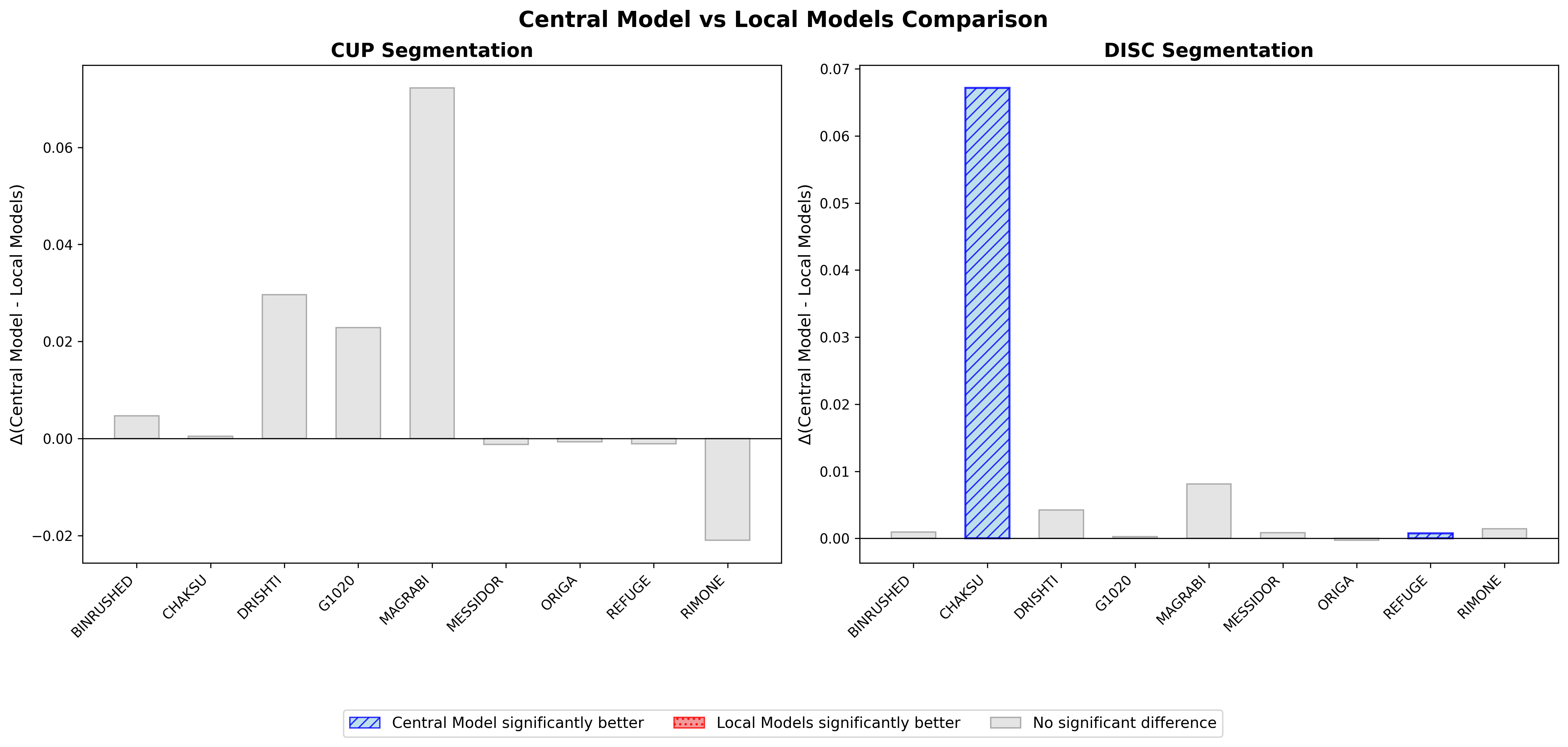

**Figure 3: Results for model generalizability comparing Global Validation vs local models.**

(a) **Bar plots showing difference of means and Wilcoxon signed-rank test significant winners for optic cup segmentation for Global Validation when compared to local models evaluated on the same dataset as well as external datasets.**

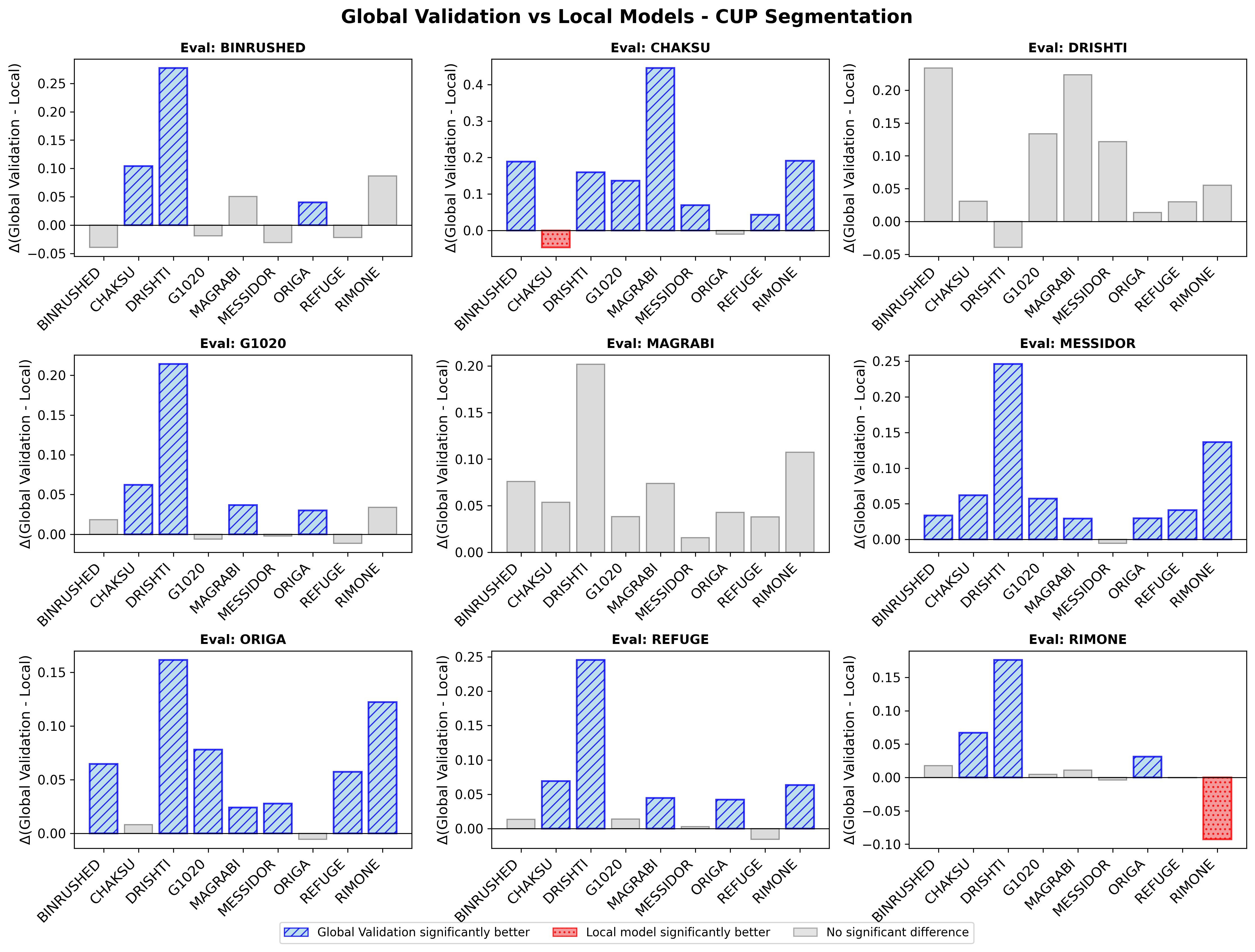

(b) **Bar plots showing difference of means and Wilcoxon signed-rank test significant winners for optic disc segmentation for Global Validation when compared to local models evaluated on the same dataset as well as external datasets.**

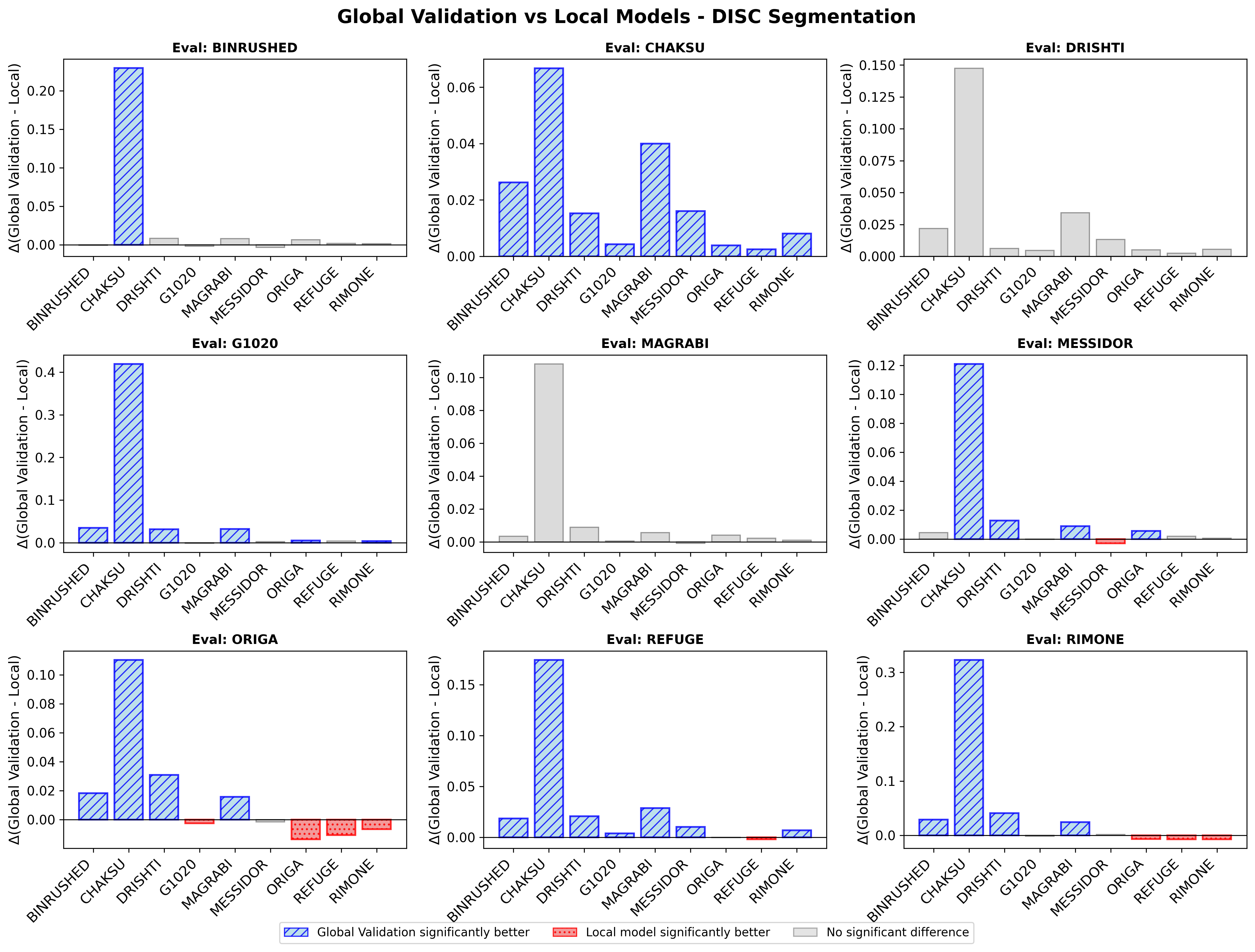

**Figure 4: Results for model generalizability comparing Weighted Global Validation vs local models.**

(a) **Bar plots showing difference of mean dice scores and Wilcoxon signed-rank test significant winners for optic cup segmentation for Weighted Global Validation when compared to local models evaluated on the same dataset as well as external datasets.**
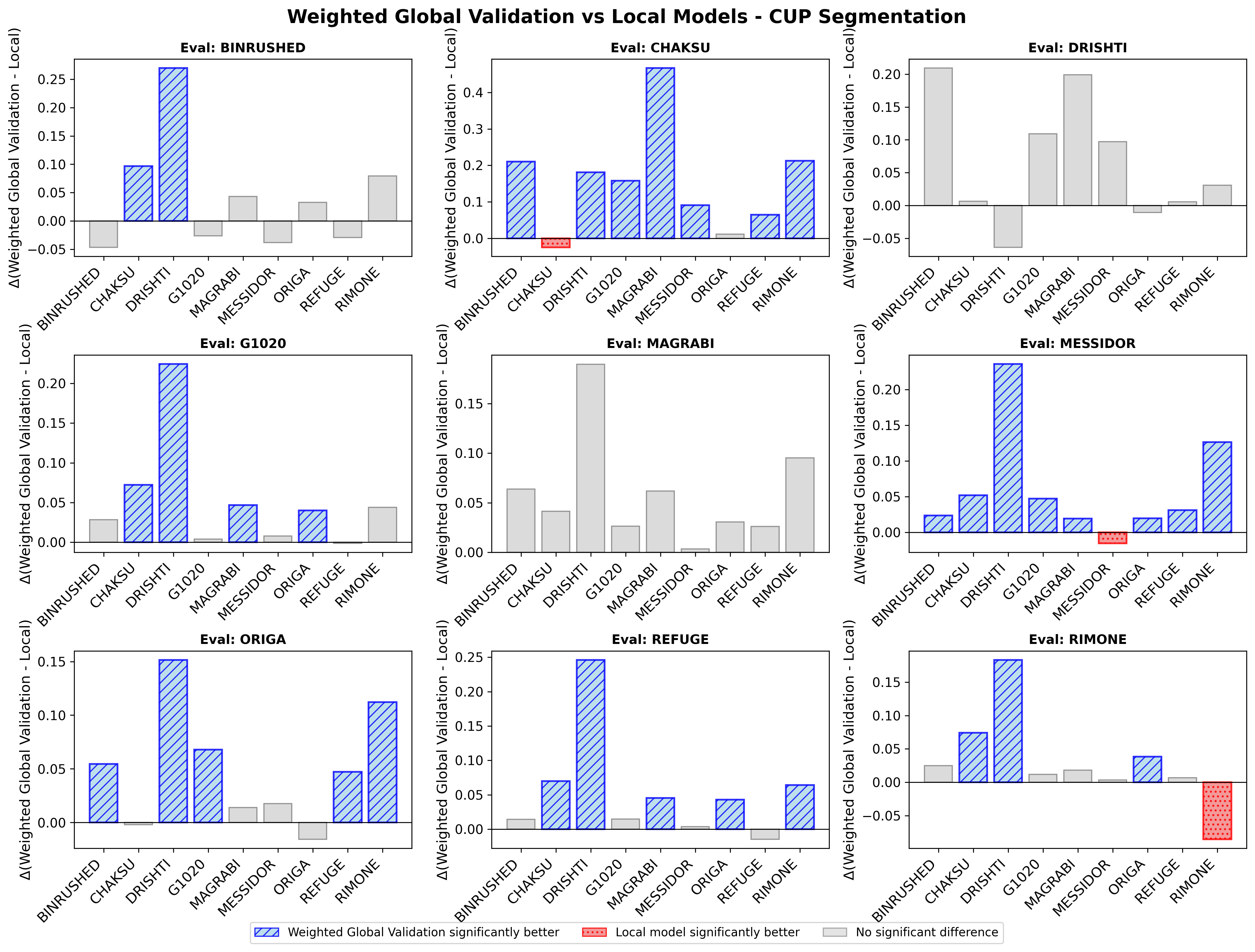

(b) **Bar plots showing difference of mean dice scores and Wilcoxon signed-rank test significant winners for optic disc segmentation for Weighted Global Validation when compared to localmodels evaluated on the same dataset as well as external datasets.**
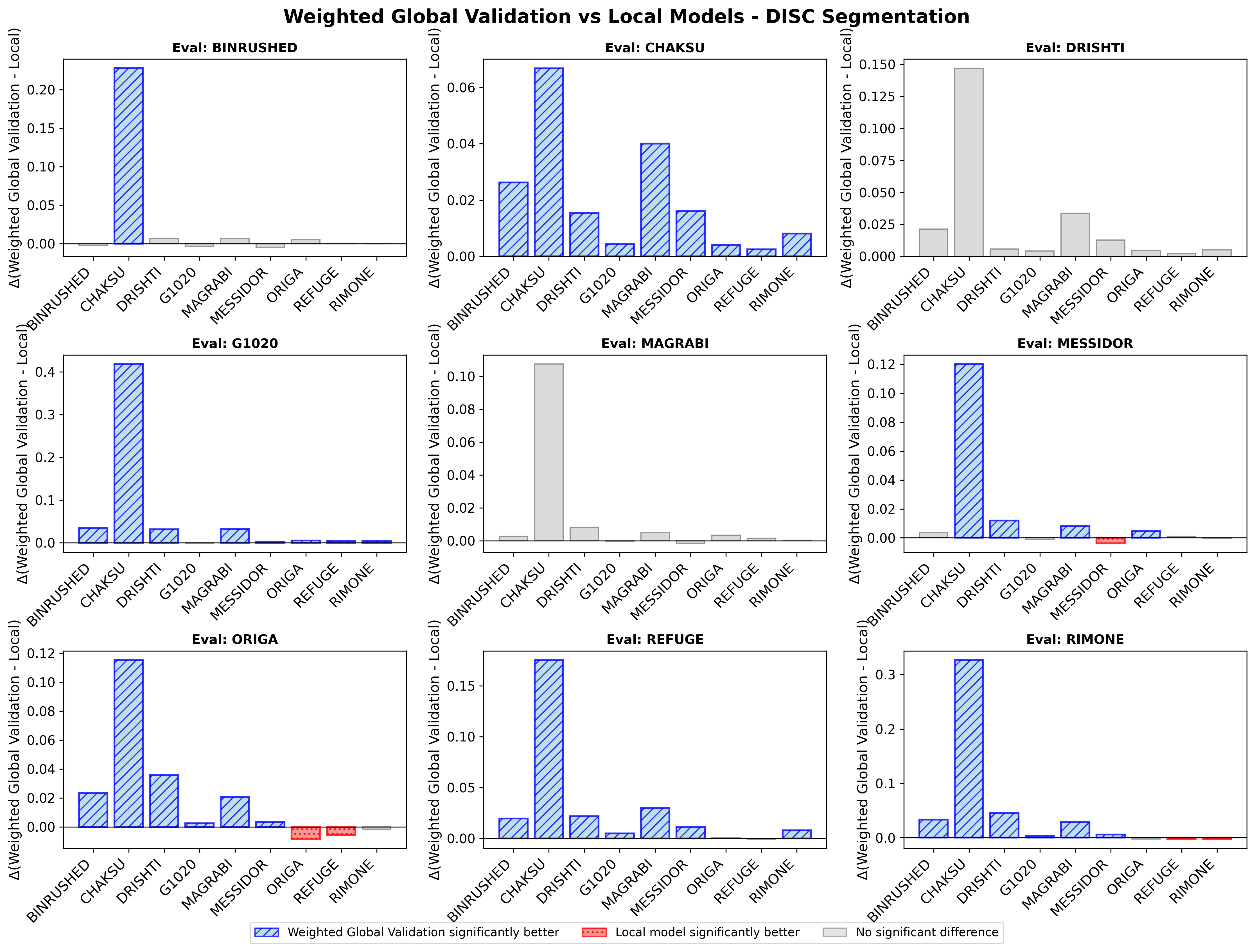

**Figure 5: Results for model generalizability comparing Onsite Validation vs localmodels.**

(a) **Bar plots showing difference of mean dice scores and Wilcoxon signed-rank test significant winners for optic cup segmentation for Onsite Validation when compared to local models evaluated on the same dataset as well as external datasets.**
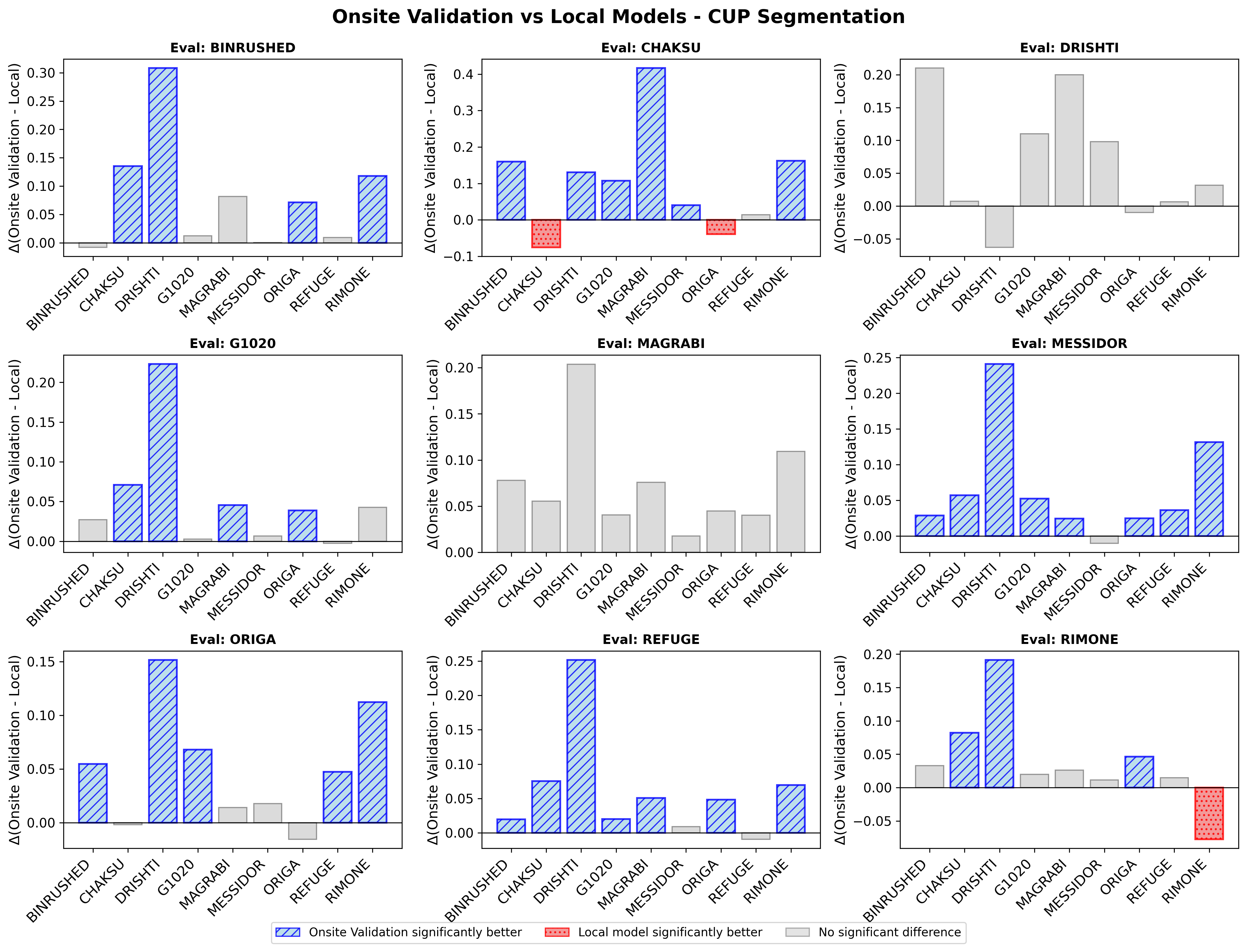

(b) **Bar plots showing difference of mean dice scores and Wilcoxon signed-rank test significant winners for optic disc segmentation for Onsite Validation when compared to local models evaluated on the same dataset as well as external datasets.**

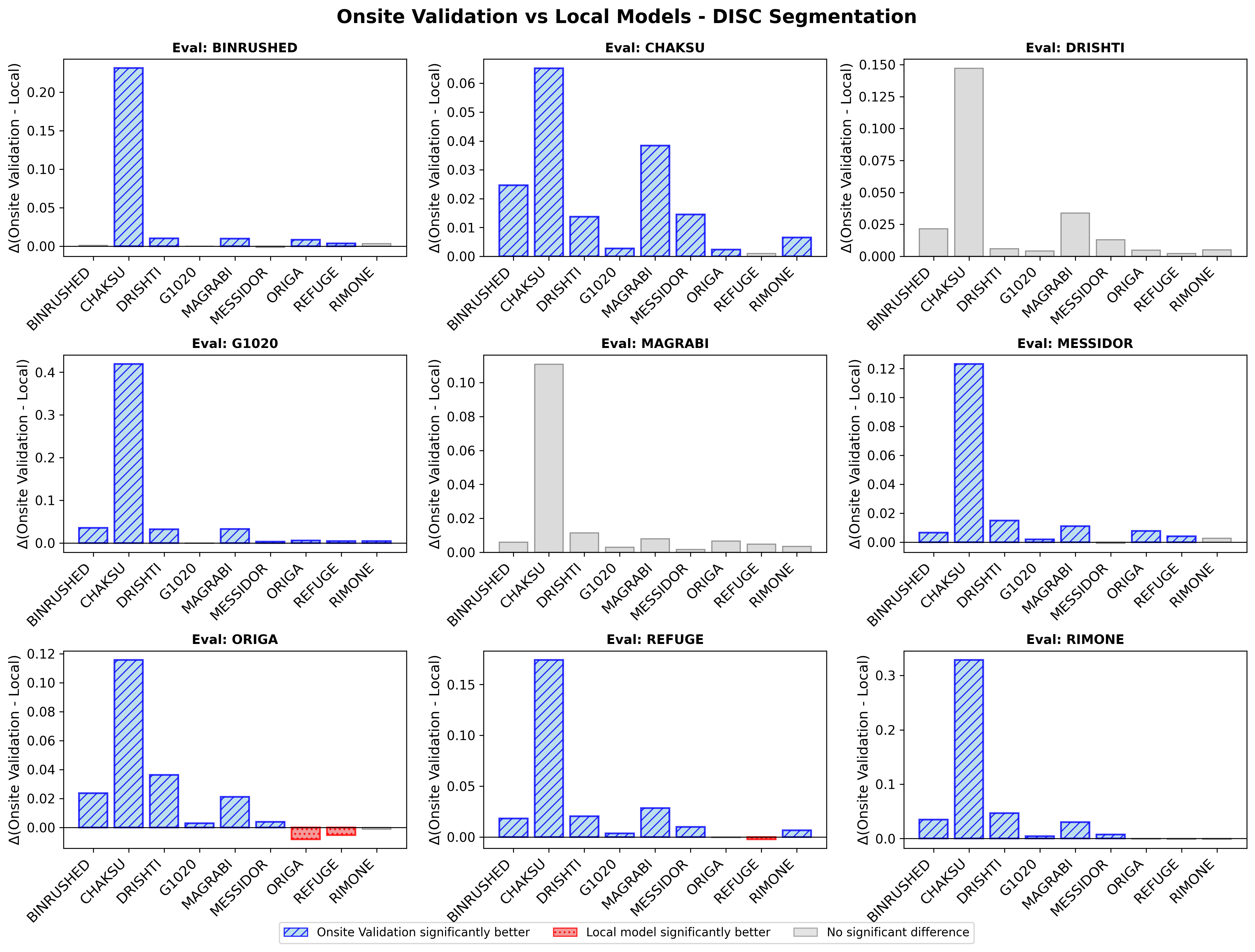

**Figure 6: Results for model generalizability comparing Central model vs local models.**

(a) **Bar plots showing difference of mean dice scores and Wilcoxon signed-rank test significant winners for optic cup segmentation for Central model when compared to local models evaluated on the same dataset as well as external datasets.**

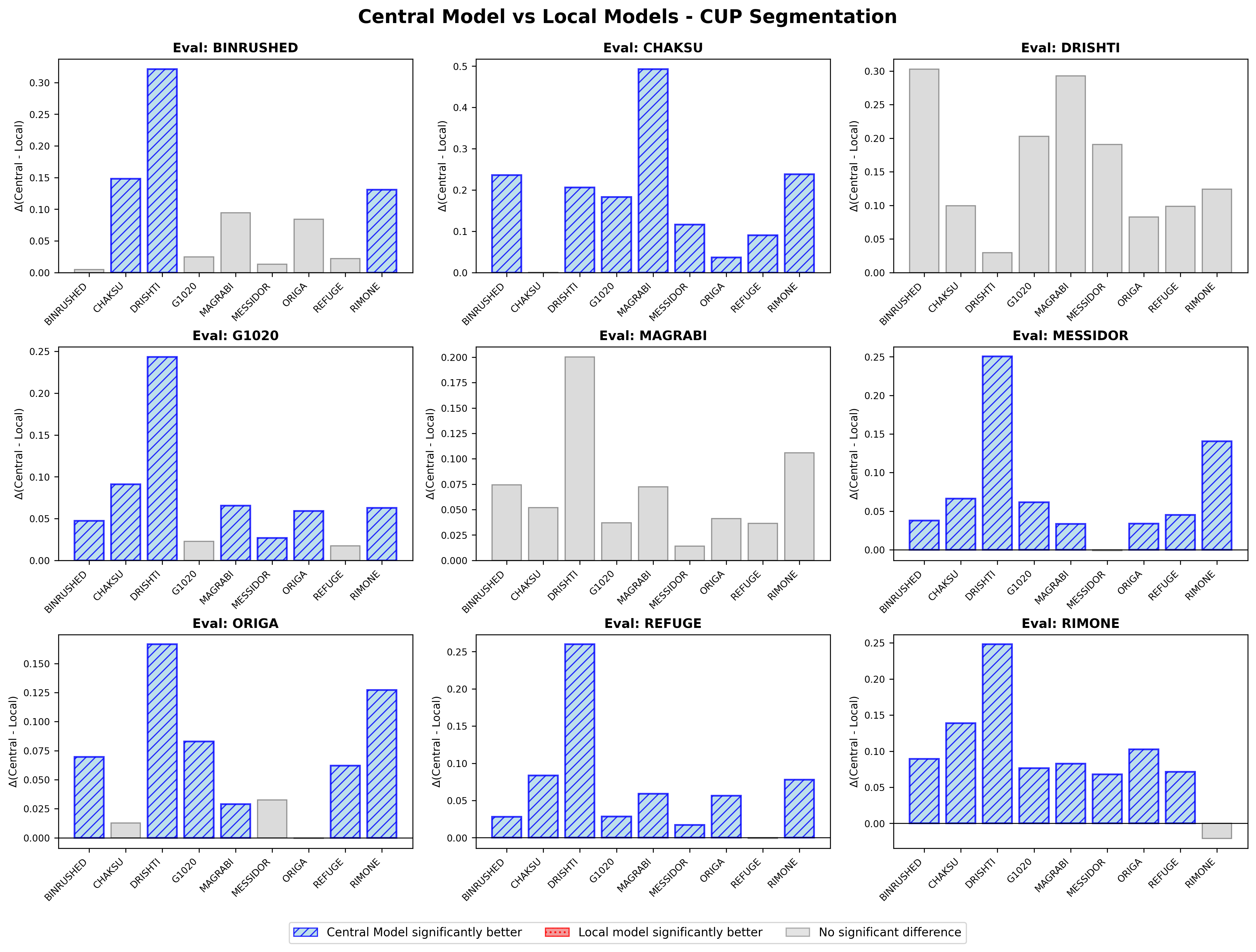

(b) **Bar plots showing difference of mean dice scores and Wilcoxon signed-rank test significant winners for optic disc segmentation for Central model when compared to local model evaluated on the same dataset as well as external datasets.**

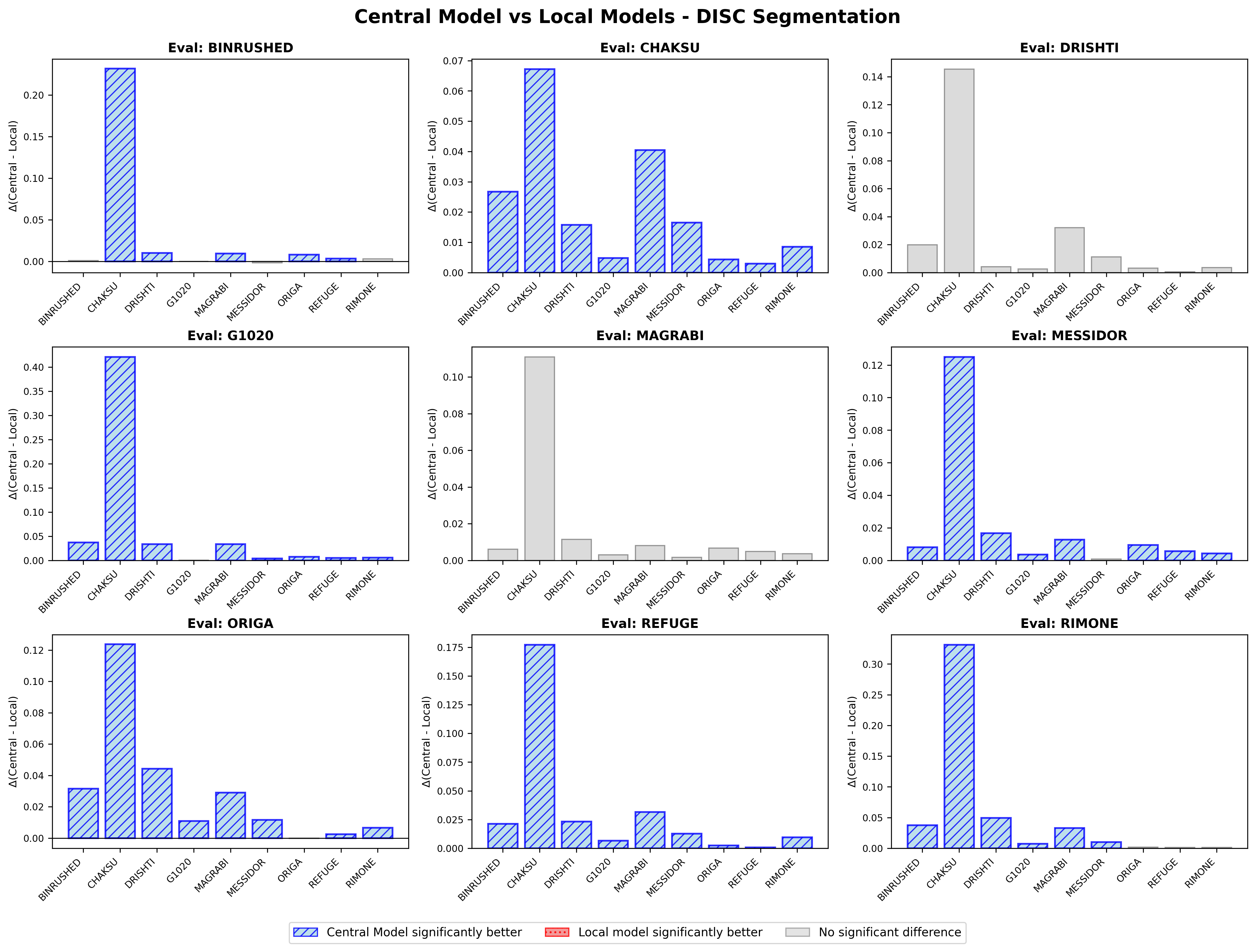

**Table 4: Results for model generalizability with total significant wins per model.**

**a) Table showing the number of times each model significantly outperformed a localmodel for optic disc segmentation, when evaluated on external datasets (different from the dataset on which the local model was trained).**

**b) Table showing the number of times each model significantly outperformed a local model for optic cup segmentation, when evaluated on external datasets (different from the dataset on which the local model was trained).**

**c) Models ranked by total significant wins over local model for both Optic disc and cup segmentation, when evaluated on external sites. As expected, central model is the best performing model, followed by Onsite Validation, Weighted Global Validation weighted and Global Validation.**

**Each column represents a specific site, and rows indicate the corresponding model. Note for Fine-Tuned Onsite Validation: While each Fine-tuned Onsite Validation model was compared against all localmodels, ideally, we need comparison with only a specific site’s localmodel (i.e. Fine-tuned Onsite Validation [binrushed] compared to localmodel trained on binrushed).**

**a)**
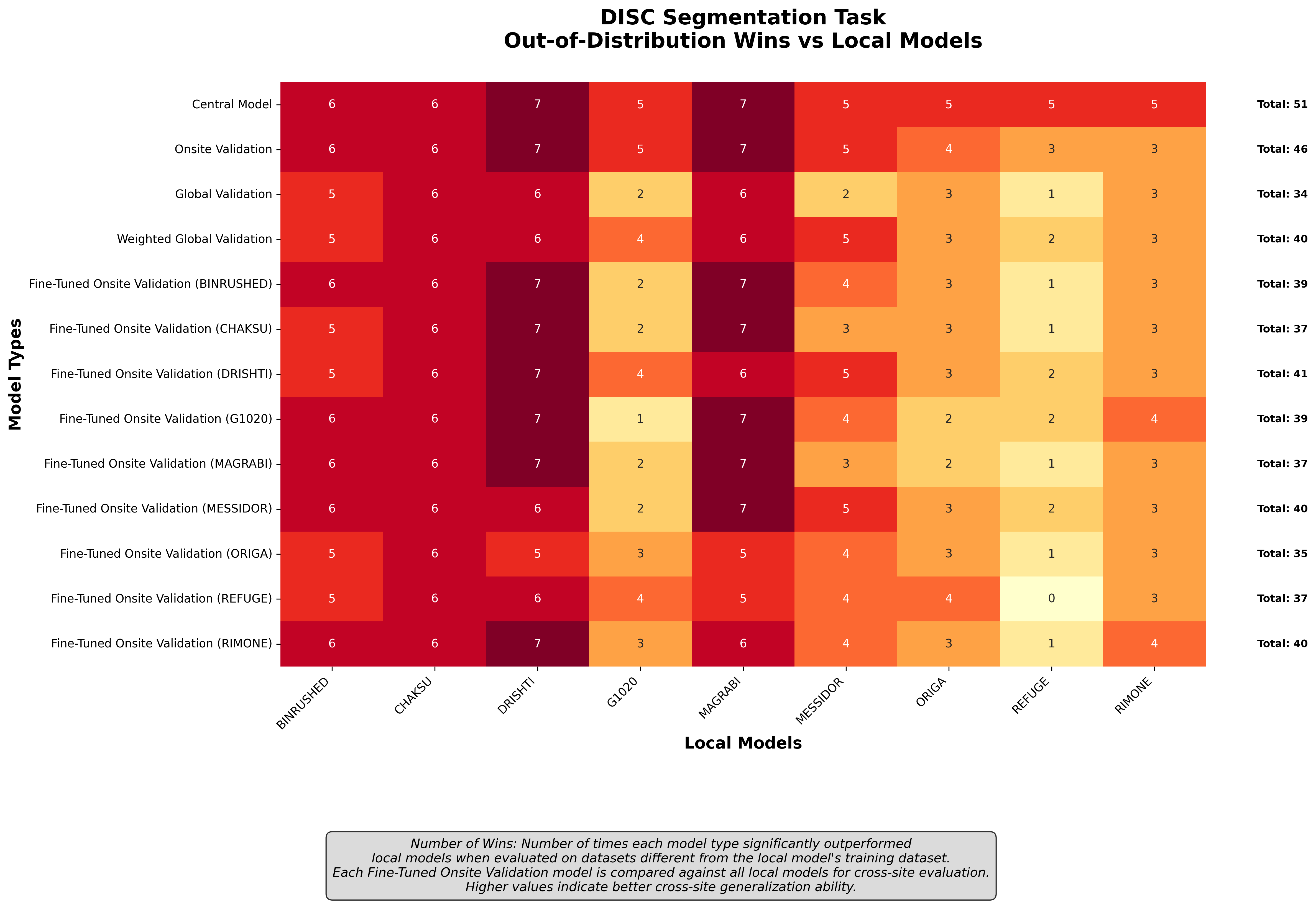

**b)**
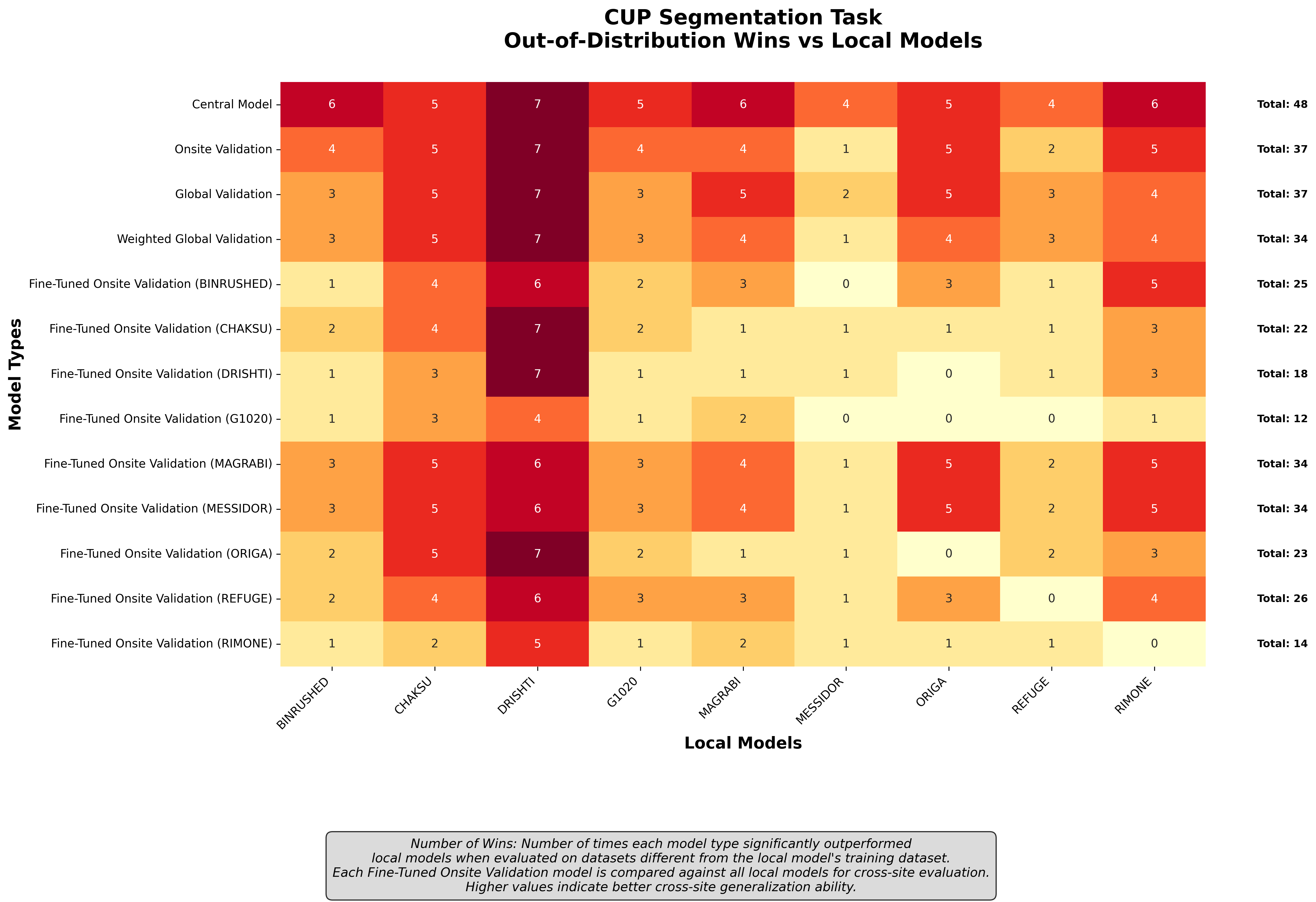

**c)**
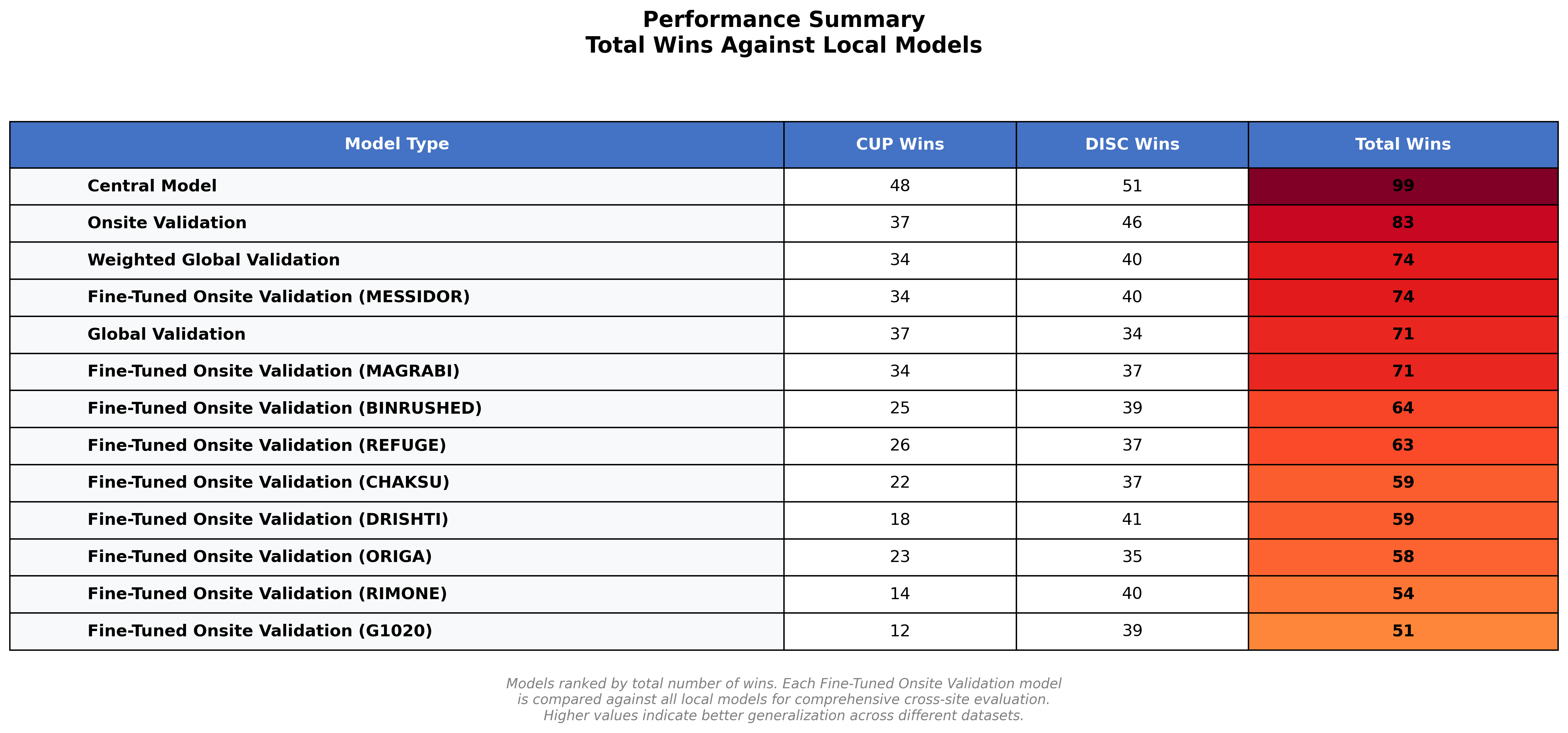

**TRIPOD-AI reporting guidelines:** A duly completed TRIPOD-AI reporting guidelines checklist (<https://www.bmj.com/content/385/bmj-2023-078378>) can be found underneath.

| Item | Description | Reported | Notes |
| --- | --- | --- | --- |
| 1 | Identify the study as developing or evaluating a multivariable prediction model, the target population, and the outcome to be predicted | Yes | Title, Abstract, Introduction |
| 2 | See TRIPOD-AI for Abstracts checklist | Partially (p.1) | The abstract incorporates several elements aligned with the TRIPOD-AI reporting structure, albeit only partially. |
| 3a | Explain the healthcare context and rationale for developing or evaluating the model | Yes | Introduction |
| 3b | Describe the target population and intended purpose of the model | Yes | Introduction, Methods |
| 3c | Describe any known health inequalities between sociodemographic groups | No | Not provided by the sources of the datasets |
| 4 | Specify the study objectives | Yes | Abstract, Introduction |
| 5a | Describe the sources of data | Yes | Methods – Datasets section |
| 5b | Specify the dates of the collected participant data | Partially | Supplementary Table 1 |
| 6a | Specify key elements of the study setting | Partially | Supplementary Table 1 |
| 6b | Describe the eligibility criteria for study participants | No | Not provided by the sources of the datasets |
| 6 | Give details of treatments received and how handled | No | Not provided by the sources of the datasets |
| 7 | Describe any data pre-processing and quality checking | Yes | Supplementary: Image Preprocessing |
| 8a | Define the outcome being predicted | Yes | Optic Disc and Cup Segmentation provided in Introduction |
| 8b | If outcome assessment requires subjective interpretation, describe qualifications | Yes | Supplementary Table 1 |
| 8c | Report any actions to blind assessment | No | Not reported |
| 9a | Describe choice of initial predictors | Yes | Color fundus photos. |
| 9b | Clearly define all predictors | Yes | Defined in Methods under Datasets. |
| 9c | If predictor requires subjective interpretation, describe assessor qualifications | Yes | Supplementary Table 1. |
| 10 | Explain how the study size was arrived at | No | Not provided by the sources of the datasets |
| 11 | Describe how missing data were handled | No | Not provided by the sources of the datasets |
| 12a | Describe how data were used (partitioning, internal validation, etc.) | Yes | 80/10/10 splits, train/val/test process |
| 12b | Describe how predictors were handled | Yes | Reported in Supplementary Image pre-processing. |
| 12c | Specify type of model and model-building steps | Yes | All model training details provided in Methods |
| 12d | Describe how heterogeneity in estimates was handled | Yes | Bonferroni Correction for multiple hypothesis testing at an alpha value of 0.05 was used for p-value correction |
| 12e | Specify all measures and plots to evaluate model performance. | Yes | Sørensen–Dice coefficient (or Dice score) and Wilcoxon signed rank test for model comparisons. |
| 12f | Describe model updating (e.g., recalibration) | No | Not performed due to limited dataset size |
| 12g | For model evaluation, describe how predictions were calculated | Yes | Outlined in supplementary under Model architecture and Loss functions |
| 13 | If class imbalance methods used, describe how | N/A |  |
| 14 | Describe any approaches used to address model fairness | No | Constrained by incomplete demographic information, which limits the ability to evaluate model fairness. |
| 15 | Specify the output of the prediction model and rationale | Yes | OD and OC segmentations. |
| 16 | Identify any differences between development and evaluation data | No | Performed an 80/10/10 splits into train/val/test |
| 17 | Name the institutional review board or ethics committee | Yes | Listed for all institutions in Methods. |
| 18a | Give source of funding and funder's role | Yes | NIH and other funding detailed under Acknowledgements. |
| 18b | Declare any conflicts of interest | Yes | Conflict of Interest section |
| 18c | State whether study protocol can be accessed | No | Not provided by the sources of the datasets |
| 18d | Provide registration info or state not registered | No | Not provided by the sources of the datasets |
| 18e | Provide availability of the study data | Yes | Sources of data provided in Supplementary |
| 18f | Provide availability of the analytical code | Yes | Github link provided for all analytical code. |
| 19 | Provide details of any patient/public involvement | No | Not mentioned |
| 20a | Describe flow of participants through study | No | Not provided by the sources of the datasets |
| 20b | Report participant characteristics | Partially | Table 1 |
| 20c | Show comparison between development and test data | No | Performed an 80/10/10 splits into train/val/test |
| 21 | Specify number of participants and outcome events | Yes | Throughout Results and Tables (including Supplementary tables) |
| 22 | Provide details of the full prediction model | Yes | Described in Methods, weights shared on hugging face. |
| 23a | Report model performance estimates with confidence intervals | Partially | Dice score reported. CI not reported due to limitations on the number of experiments. |
| 23b | If examined, report heterogeneity in performance | No | CI not reported due to limitations on the number of experiments. |
| 24 | Report results from any model updating | No | Model updating not done |
| 25 | Give overall interpretation of the main results | Yes | Discussion |
| 26 | Discuss any limitations | Yes | Detailed in Future Work/Limitations |
| 27a | Describe how poor/unavailable input data were handled | No | Not provided by source of datasets |
| 27b | Specify requirements for handling input model data | Yes | Listed in Supplementary under image pre-processing. |
| 27c | Discuss any next steps for future research | Yes | Detailed in Future Work/Limitations |
